# Development of a Risk Prediction Model for Sepsis-Related Delirium Based on Multiple Machine Learning Approaches and an Online Calculator

**DOI:** 10.1101/2025.04.17.25325998

**Authors:** Lang Gao, Guang Dong Wang, Xing Yi Yang, Shi Jun Tong, Xu Jie Wang, Yun Ruo Chen, Jin Ying Bai, Ya Xin Zhang

**Affiliations:** Department of Critical Care Medicine, Clinical Medical College of Qinghai University, Xi ning, 810000,China; Department of Respiratory and Critical Care Medicine, First Affiliated Hospital of Xi’an Jiaotong University, Xi’an, Shanxi, 710061, China; Department of Gastroenterology Disease, XianJu People’s Hospital, Zhejiang Southeast Campus of Zhejiang Provincial People’s Hospital. Affiliated Xianju’s Hospital, Hang zhou Medical College, Xianju Zhejiang, 318000,China; Department of Emergency Medicine, Clinical Medical College of Qinghai University, Xi ning, 810000,China; Department of Neurology, Xia men Humanity Hospital, Fujian Medical University, Xia men Fujian, 361009, China

**Author notes:** **Corresponding author : Email:****(YXZ)**.

**Keywords:** sepsis, delirium, machine learning, gradient boosting machine, online calculator

## Abstract

**Background:** Sepsis-associated delirium (SAD) occurs due to disruptions in neurotransmission linked to inflammatory responses from infections. It poses significant challenges in clinical management and is associated with poor outcomes. Survivors often experience long-term cognitive and behavioral issues that impact their quality of life and place a burden on their families. This study aims to create and validate an interpretable machine learning algorithm for predicting SAD. We will also develop an online web calculator based on this model to assist in assessing the risk of SAD onset in clinical settings.

**Methods:** This study is a retrospective analysis utilizing data from 16120 patients in the Medical Information Mart for Intensive Care IV database. To manage imbalanced data, we applied the SMOTE method. Feature selection was conducted using Multivariate Logistic Regression, Lasso regression, and the Boruta algorithm. We developed predictive models using eight machine learning algorithms and selected the best one for validation. The SHapley Additive exPlanations (SHAP) method was used for visualization and interpretation, enhancing the clinical understanding of the model, alongside the creation of an online web calculator.

**Results:** We combined three feature selection methods to identify 17 key features for our machine learning prediction model. The Gradient Boosting Machine showed strong calibration and excellent predictive performance in the internal validation set. The SHAP feature importance ranking revealed five critical risk factors for predicting outcomes: Glasgow Coma Scale, ICU Day, Chloride, Sodium, and Sequential Organ Failure Assessment. Based on this optimal model, we successfully developed an online web calculator.

**Conclusion:** We developed a machine learning predictive model for SAD that showed strong performance and clinical utility in accurately predicting SAD. Additionally, we created a user-friendly web calculator to help healthcare professionals identify SAD early, allowing for timely treatment adjustments and improved patient outcomes.

## 1. Introduction

Sepsis is a host’s uncontrolled systemic inflammatory response syndrome triggered by infection, where a large number of inflammatory factors lead to multi-organ dysfunction, ultimately resulting in a crisis of multiple organ failure that can threaten the patient’s life (1). Although supportive bundled care for sepsis has been helpful in reducing mortality rates among sepsis patients, the overall prognosis remains poor. According to the Centers for Disease Control and Prevention, the annual incidence of sepsis in the United States is 300-1,000 cases per 100,000 people. Global burden of disease studies show that the global annual incidence of sepsis reaches up to 31 million cases, making it a major public health threat (2–4).

Delirium is an acute brain dysfunction characterized by fluctuations in attention and impaired cognitive function, with diverse symptoms that may include psychomotor agitation and altered consciousness. It is commonly seen in critically ill patients admitted to the intensive care unit (ICU) (5). Sepsis is one of the major risk factors for delirium, as the systemic inflammatory response syndrome associated with sepsis can disrupt the balance of central nervous system function, leading to delirium in patients. Delirium in sepsis patients is referred to as SAD; however, the mechanisms by which sepsis affects the central nervous system remain unclear and may involve brain inflammation, cerebral perfusion, blood-brain barrier disruption, and neurotransmitter disturbances. Managing SAD in the ICU has historically been challenging, with poor prognoses for SAD patients. Survivors often experience long-term and severe cognitive impairments and behavioral abnormalities, significantly reducing their quality of life and placing a heavy burden on families (6, 7). Therefore, early identification and prevention of potential SAD patients in clinical practice are of utmost importance. Consequently, we developed a predictive model for SAD and constructed an online calculator to provide clinicians with an important tool for early identification of high-risk populations, optimizing individual intervention measures through various simple clinical indicators to improve clinical outcomes and reduce the length of hospital stay.

The rapid development of artificial intelligence and machine learning algorithms has significantly accelerated the innovation of predictive models for medical diagnosis and prognosis assessment of various diseases (8, 9). Compared to traditional regression analysis, machine learning is widely used to handle clinical data, extracting features related to clinical outcomes from large datasets and identifying independent predictive factors associated with clinical results, thereby better addressing clinical decision-making issues (10, 11). In the ICU, assessing a patient’s risk of developing delirium heavily relies on subjective judgment by healthcare professionals, necessitating the development of a more straightforward, objective, and accurate tool to evaluate the risk of delirium. The aim of this study is to establish an interpretable machine learning-based online predictive tool for SAD risk to assist clinicians in accurately assessing the risk of delirium early, allowing timely adjustments to treatment plans to reduce the incidence of delirium and make more informed clinical decisions.

## 2. Materials and methods

### 2.1 Study design

The data for this study were sourced from the Medical Information Mart for Intensive Care IV-3.1 (MIMIC-IV 3.1) database. The MIMIC-IV 3.1 database is a multicenter critical care clinical database developed by the MIT Laboratory for Computational Physiology, and it is one of the largest and most comprehensive medical data resources of its kind globally. This database contains comprehensive information on nearly 95,000 patients from the Beth Israel Deaconess Medical Center between 2008 and 2022, including basic demographic information, diagnostic and medication data, laboratory test results, nursing records, and outcome data (12, 13). To utilize the MIMIC-IV 3.1 database, we completed the web-based course provided by the National Institutes of Health (NIH), fulfilling the CITI program training requirements and obtaining research ethics certification (certification number: 66380198). Our study complies with the Declaration of Helsinki and international medical ethics standards, and all patient data were fully anonymized to waive informed consent requirements and ethical review.

### 2.2 Study population and outcome

We extracted each patient’s hospitalization information, including demographics, vital signs, laboratory test indicators, and treatment information, from three databases using Structured Query Language (SQL). Patients included in the study met the following criteria: (1) confirmed or suspected infection within 24 hours of ICU admission, and according to the Sepsis-3.0 diagnostic criteria, a Sequential Organ Failure Assessment (SOFA) score of ≥ 2. (2) Patients were aged 18-100 years and had laboratory test records within 24 hours of ICU admission. (3) first admission to the ICU. (4) Patients assessed using the Confusion Assessment Method for the Intensive Care Unit (CAM-ICU). CAM-ICU is an effective screening tool for identifying delirium in the intensive care unit, and septic patients with positive assessments are defined as having SAD. The exclusion criteria were as follows: (1) patients with dementia or schizophrenia; (2) missing variables greater than 30%; (3) ICU stay of less than 24 hours; (4) outliers in vital signs. (5) develop delirium before admission to the ICU. The data collection process for the training and testing sets is illustrated in Fig 1.

**Fig 1.**
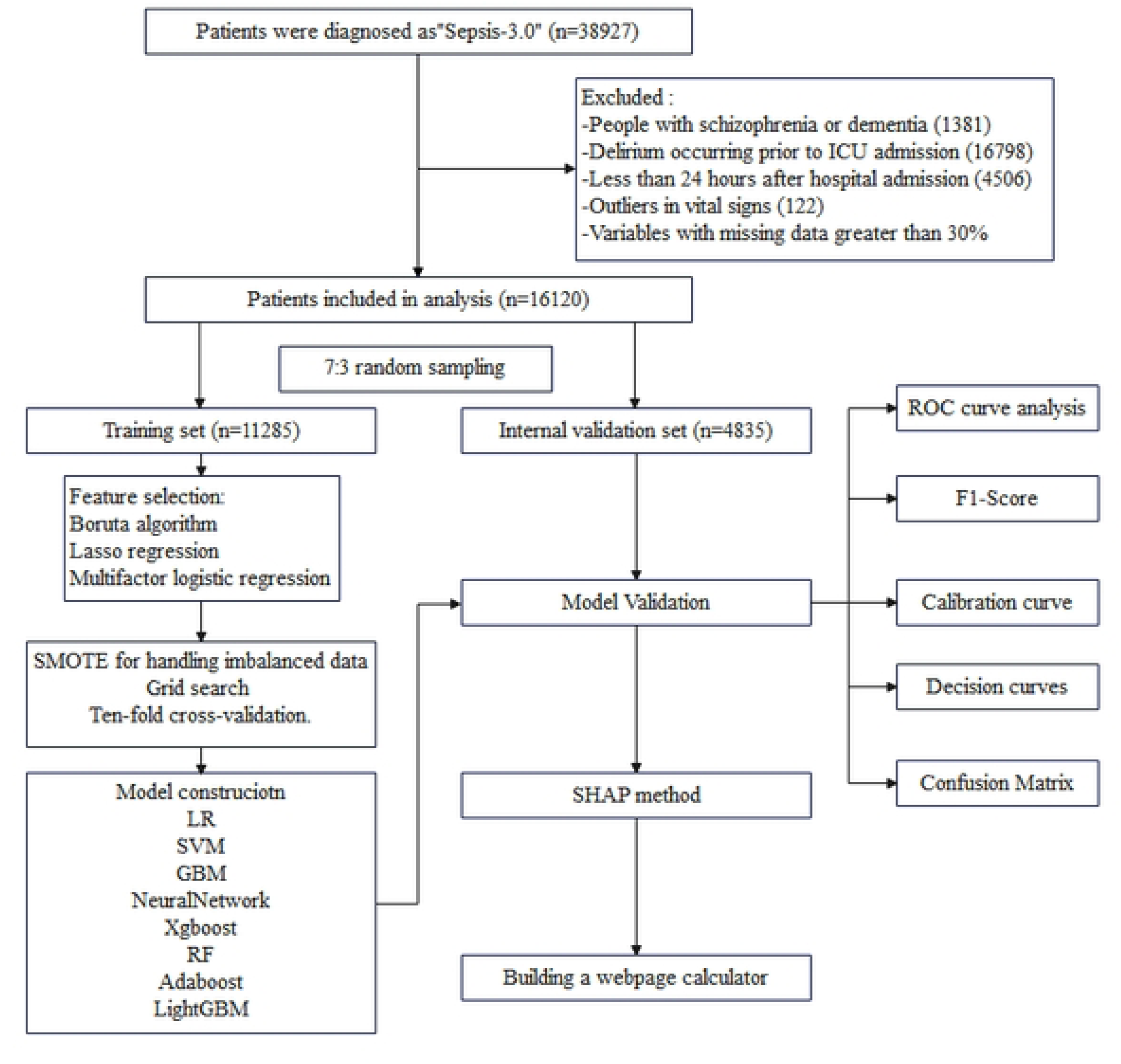
**The entire research flowchart.**

### 2.3 Inclusion of variables

This study retrospectively collected clinical data from patients, encompassing 46 variables. The final dataset included demographic characteristics and baseline clinical information, such as Age, Weight, Gender, and ICU Day; vital signs within 24 hours of ICU admission, including HR, SBP, DBP, MAP, RR, SPO2, Temperature, SOFA, Simplified Acute Physiology Score II (SAPS II), and Glasgow Coma Scale (GCS); laboratory test data obtained within 24 hours of ICU admission, such as Red Blood Cell count, White Blood Cell count, Platelet count, Hemoglobin, Red Cell Distribution Width (RDW), Hematocrit, Mean Corpuscular Hemoglobin (MCH), Mean Corpuscular Hemoglobin Concentration (MCHC), Mean Corpuscular Volume (MCV), International Normalized Ratio, Prothrombin Time, Activated Partial Thromboplastin Time, Creatinine, Blood Urea Nitrogen, Anion Gap, Potential of Hydrogen (PH), Bicarbonate, Calcium, Magnesium, Chloride, Potassium, Sodium, Lactate, and Glucose; along with certain comorbidities such as Hypertension, Acute Kidney Injury (AKI), Type 2 Diabetes Mellitus, and Heart Failure. Iatrogenic factors and environmental factors included various interventions received by patients during their ICU stay, which encompassed pharmacological treatments and organ support techniques, such as Mechanical Ventilation (MV) and Continuous Renal Replacement Therapy (CRRT), as well as medications like Midazolam and Vasopressin (VP). We employed multiple imputation methods to address missing data.

### 2.4 The Selection of Features and the Processing of Data

We randomly divided the included patients into a training set (70%) and a validation set (30%) using random sampling. In the training set, there were 2,355 SAD cases and 8,930 Non-SAD cases. The imbalance in sample size could lead to misleading results from the machine learning predictive model. To address this, we used the SMOTE algorithm to balance the training set, resulting in 4,710 SAD and 4,710 Non-SAD cases. This approach alleviates the bias introduced by sample imbalance while avoiding the risks associated with excessive oversampling. The training set was used for model development, while the validation set was employed to assess the model’s performance. To determine potential predictive factors in the training set, we utilized three independent methods to filter baseline variables: Multiple Linear Regression(MLR), Least Absolute Shrinkage and Selection Operator(Lasso), and the Boruta algorithm (14). MLR is a variable selection method based on covariate adjustment that controls for confounding factors. By utilizing statistical significance (P < 0.05), MLR identifies variables independently associated with outcomes while retaining predictive factors that remain interpretable in the presence of multivariable co-occurrence, providing adjusted odds ratios (OR) and their confidence intervals. This process results in a validated variable set with both statistical and clinical significance, offering robust support for clinical decision-making (15). Lasso regression, when handling high-dimensional data, applies penalties to feature coefficients, automatically selecting variables that have a practical impact on the prediction target. When multiple features are correlated, Lasso regression retains the most representative feature while eliminating multicollinearity interference (16, 17). To optimize model effectiveness, we employed 20-fold cross-validation to analyze baseline high-dimensional data and selected variables, which mitigated the risk of overfitting and ensured result stability.The Boruta algorithm is an ensemble feature selection framework based on random forests, where the core idea is to conduct significance testing on original variables by constructing “shadow features” (18). The Boruta algorithm is capable of capturing complex nonlinear relationships, effectively reducing the risk of overfitting while enhancing model performance, thus providing a more reliable feature space for subsequent model construction (19). The predictive model was constructed using the shared variables identified through MLR, Lasso regression, and the Boruta algorithm.

### 2.5 Model development and evaluation

We utilized eight machine learning methods to construct models in the training set: Logistic Regression (LR), Support Vector Machine (SVM), Gradient Boosting Machine (GBM), Neural Network, Random Forest (RF), Extreme Gradient Boosting (XGBoost), Adaptive Boosting (AdaBoost), and Light Gradient Boosting Machine (LightGBM). In this study, we employed the “caret” package to perform model selection using 10-fold cross-validation, while incorporating a grid parameter optimization process during the model training phase, as detailed in S1 Table (20). Subsequently, we tested the models’ performance using the internal validation set, evaluating different models through the Area Under the Receiver Operating Characteristic Curve (AUC), Decision Curve Analysis (DCA), and calibration curves. Additionally, we calculated the Accuracy, Sensitivity, and F1-score for each model to further assess performance. Based on the evaluation results, we selected the best-performing model for SHAP significance analysis and generated SHAP summary plots to assess feature importance. We then conducted SHAP dependence plots to analyze the mechanisms by which features influence the prediction outcomes, and finally quantified the contribution weights of each feature within individual samples using SHAP analysis. For ease of use by clinicians, we developed a user-friendly web calculator.

### 2.6 Statistical analysis

In this study, all statistical analyses were performed using R software (version 4.4.2) and Python software (version 3.10.6). To compare baseline characteristics between the two groups in the MIMIC-IV dataset, continuous variables conforming to a normal distribution were expressed as Mean ± Standard Deviation (x ± s), while non-normally distributed continuous variables were denoted as Median (Interquartile Range) [M (IQR)]. Continuous variables were analyzed using independent samples t-test or Wilcoxon rank-sum test. Categorical variables were expressed as numbers (percentage) [n (%)], and the χ² test or Fisher’s exact test was employed for categorical variables, with a two-sided P < 0.05 indicating statistical significance. To address the issue of sample imbalance in the training set, we utilized the SMOTE algorithm based on the “DMwR” package in R. For model development, the dataset was randomly divided into a training set (70%) and an internal validation set (30%). Variable selection was performed using MLR, Lasso regression, and the Boruta algorithm, incorporating the shared variables from these three algorithms into the machine learning model. The “pROC” and “ggplot2” packages were employed to plot the ROC curve analysis and AUC values for the internal validation set, identifying the best predictive model. Finally, SHAP analysis was utilized to interpret the optimal model.

## 3. Results

### 3.1 Baseline characteristics

We collected data from 16120 sepsis patients in the MIMIC-IV database, with detailed screening processes illustrated in Fig 1. Among the included sepsis patients, 3,364 (26.4%) were diagnosed with delirium. Table 1 summarizes the characteristics of SAD and Non-SAD patients in the MIMIC-IV database, including demographics, treatment information, and laboratory test results. Overall, the SAD group had higher values for Age, ICU Day, SOFA, SAPS II, Creatinine, Blood Urea Nitrogen, Anion Gap, Glucose, and Platelet counts compared to the Non-SAD group, with statistical significance (P < 0.05). The median age of the study population was 67 years (57, 77), with a gender distribution of 9,532 (59%) males. We divided the total population into a training set (70%) and an internal validation set (30%), utilizing the training set for model development. To explore the correlations between variables, we plotted a correlation bar chart for each variable (Fig 2A) and a heatmap of the correlations between variables (Fig 2B).

**Fig 2.**
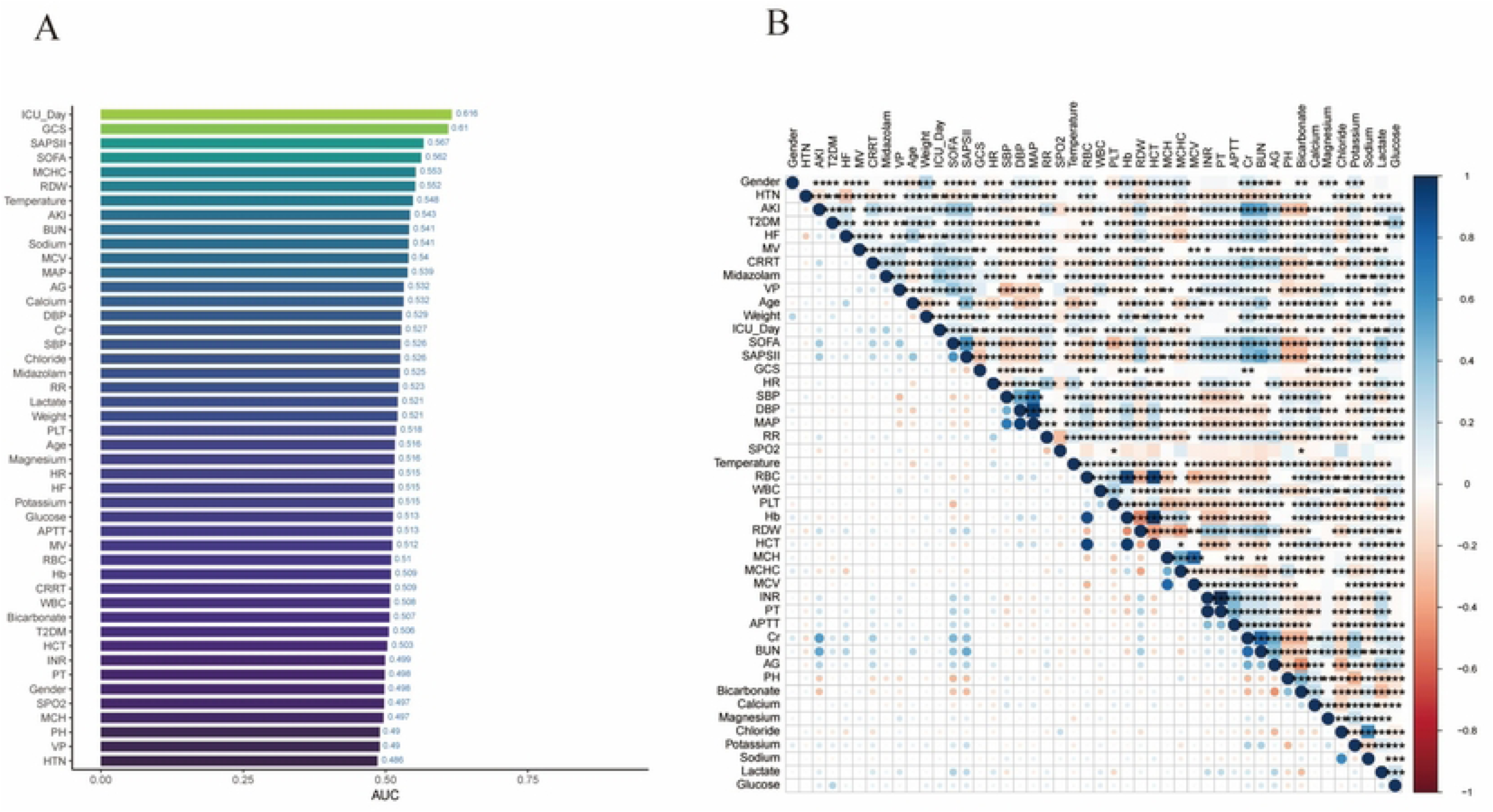
The associations between variables. (A) bar chart of variable correlation. (B) Heat map depicting the correlations among variables.

**Table 1.**
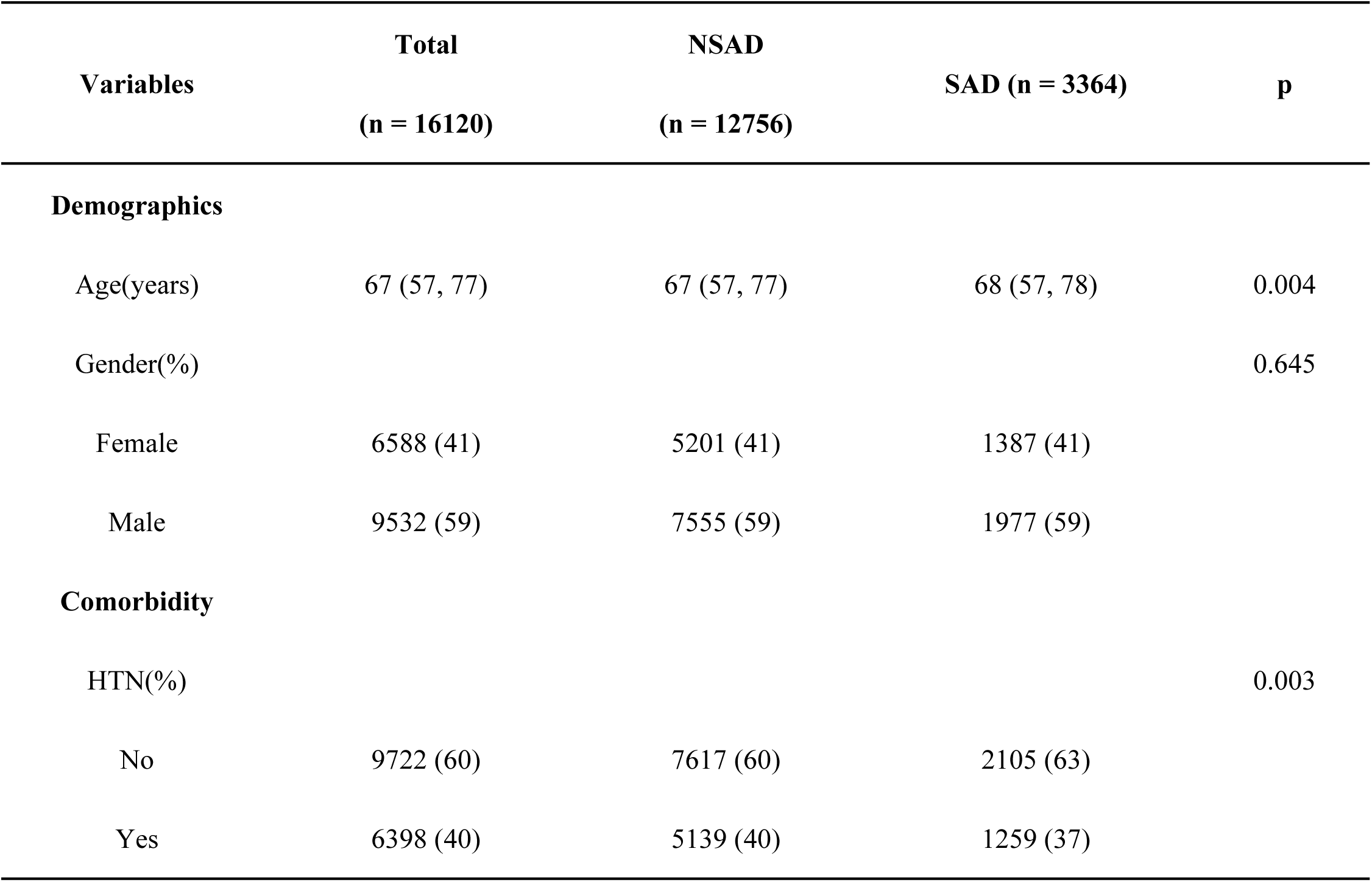
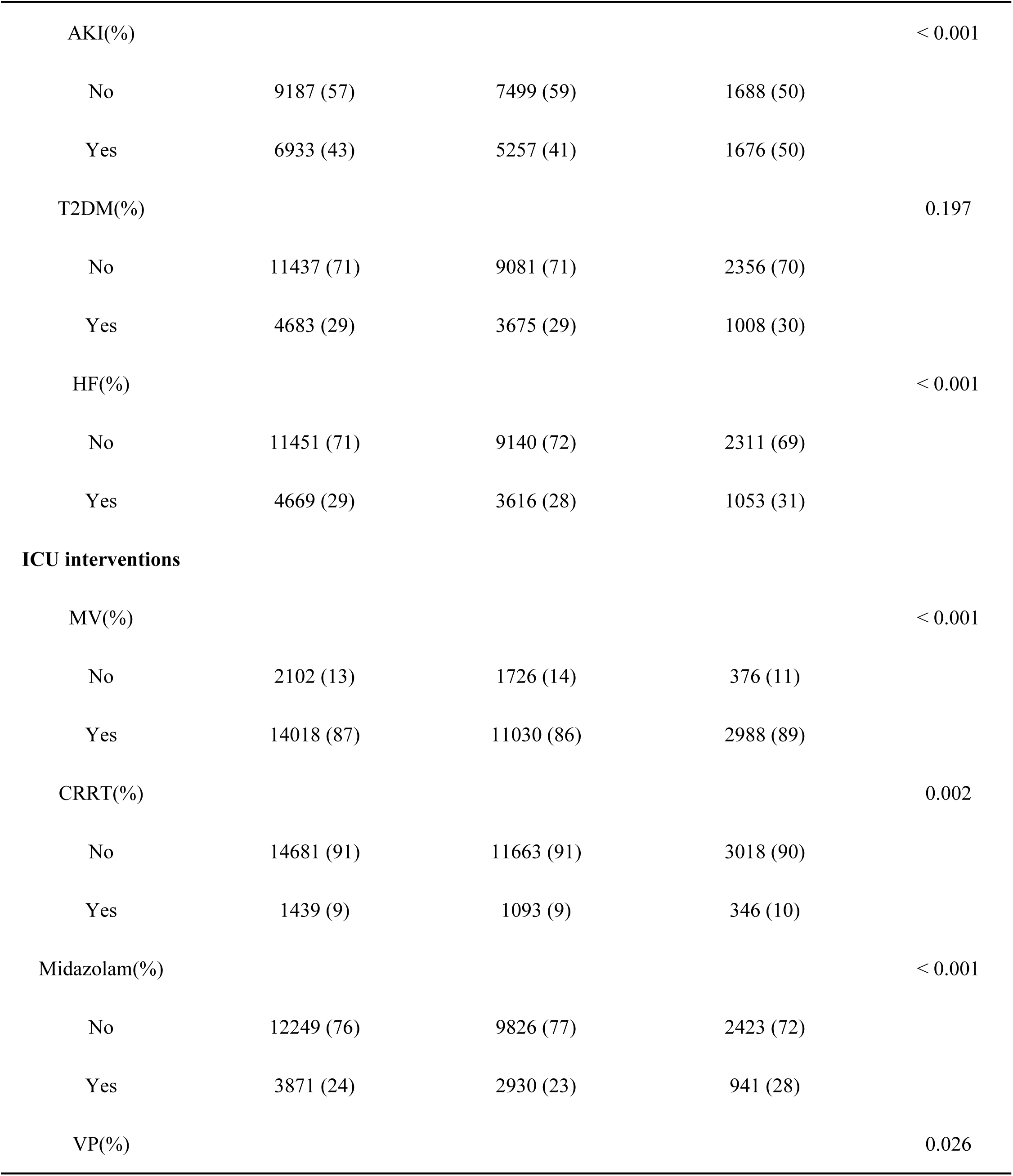
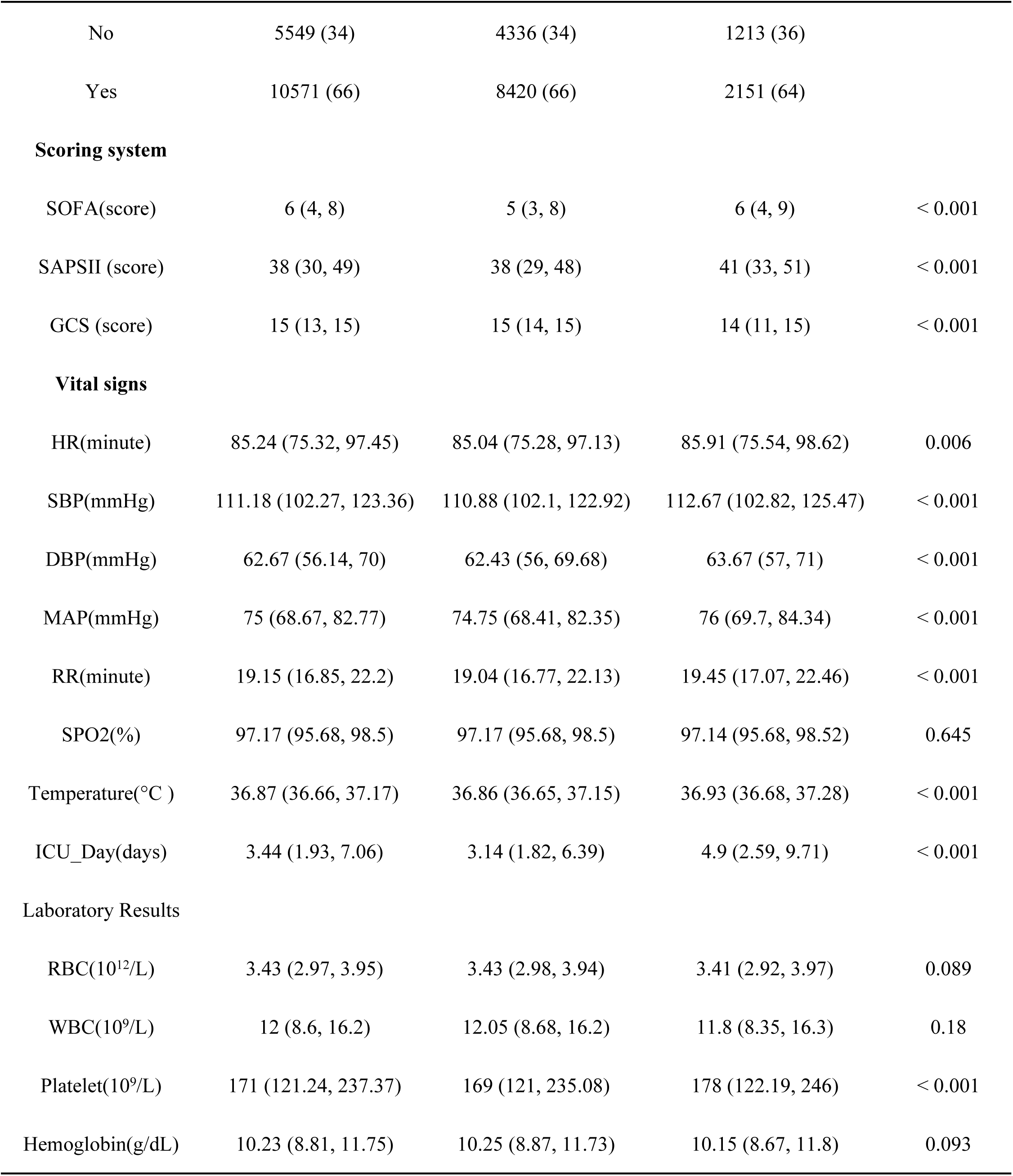
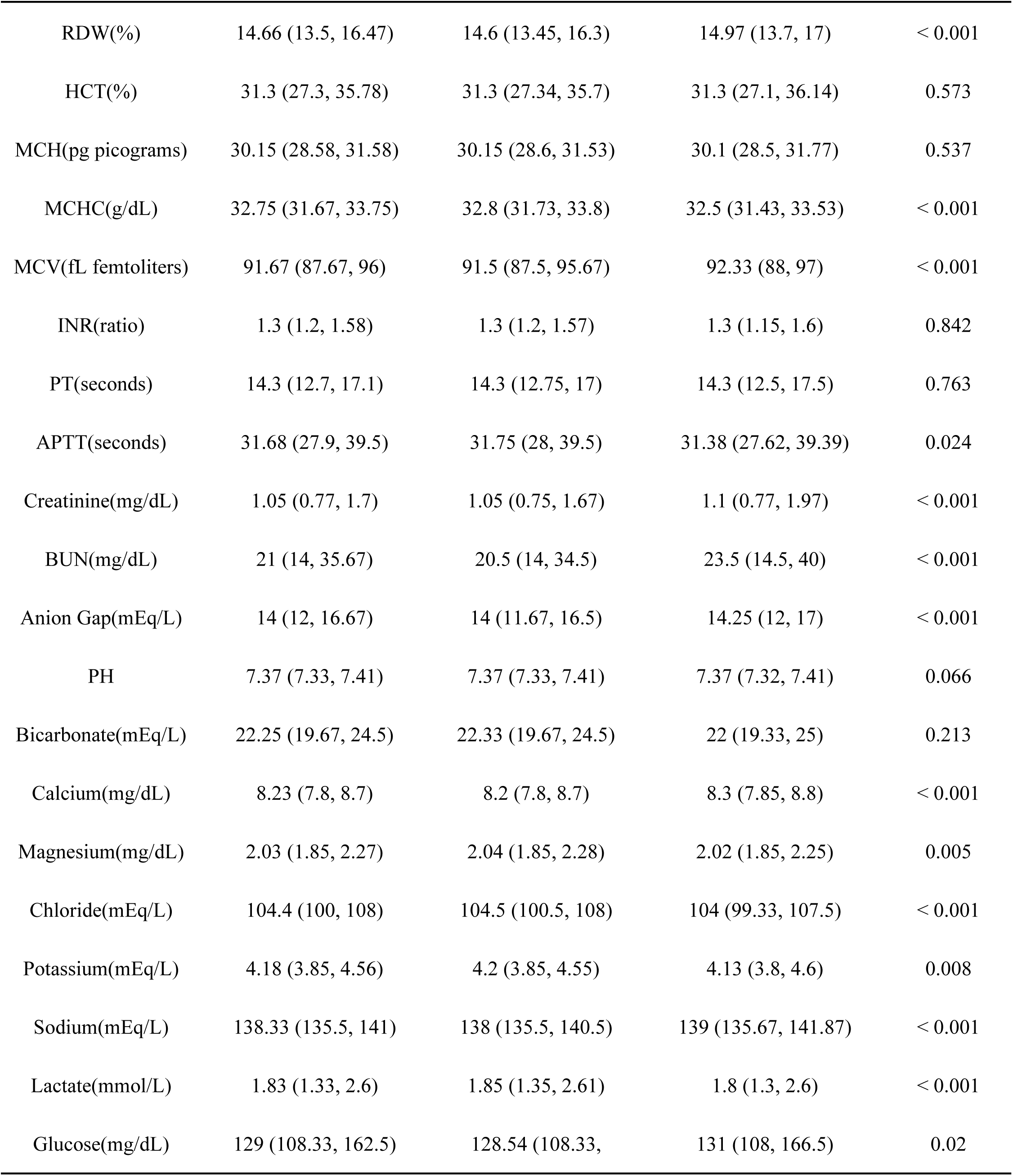
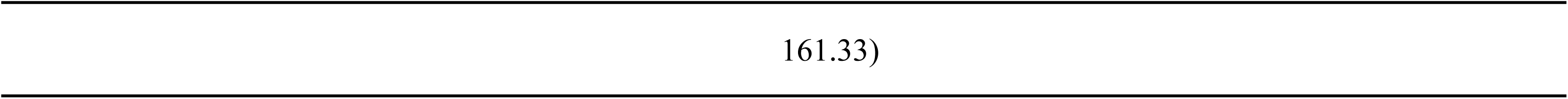
Baseline characteristics of SAD and Non-SAD patients.

| Variables | Total<br>(n = 16120) | NSAD<br>(n = 12756) | SAD (n = 3364) | p |
| --- | --- | --- | --- | --- |
| **Demographics** |  |  |  |  |
| Age(years) | 67 (57, 77) | 67 (57, 77) | 68 (57, 78) | 0.004 |
| Gender(%) |  |  |  | 0.645 |
| Female | 6588 (41) | 5201 (41) | 1387 (41) |  |
| Male | 9532 (59) | 7555 (59) | 1977 (59) |  |
| **Comorbidity** |  |  |  |  |
| HTN(%) |  |  |  | 0.003 |
| No | 9722 (60) | 7617 (60) | 2105 (63) |  |
| Yes | 6398 (40) | 5139 (40) | 1259 (37) |  |
| AKI(%) |  |  |  | < 0.001 |
| No | 9187 (57) | 7499 (59) | 1688 (50) |  |
| Yes | 6933 (43) | 5257 (41) | 1676 (50) |  |
| T2DM(%) |  |  |  | 0.197 |
| No | 11437 (71) | 9081 (71) | 2356 (70) |  |
| Yes | 4683 (29) | 3675 (29) | 1008 (30) |  |
| HF(%) |  |  |  | < 0.001 |
| No | 11451 (71) | 9140 (72) | 2311 (69) |  |
| Yes | 4669 (29) | 3616 (28) | 1053 (31) |  |
| **ICU interventions** |  |  |  |  |
| MV(%) |  |  |  | < 0.001 |
| No | 2102 (13) | 1726 (14) | 376 (11) |  |
| Yes | 14018 (87) | 11030 (86) | 2988 (89) |  |
| CRRT(%) |  |  |  | 0.002 |
| No | 14681 (91) | 11663 (91) | 3018 (90) |  |
| Yes | 1439 (9) | 1093 (9) | 346 (10) |  |
| Midazolam(%) |  |  |  | < 0.001 |
| No | 12249 (76) | 9826 (77) | 2423 (72) |  |
| Yes | 3871 (24) | 2930 (23) | 941 (28) |  |
| VP(%) |  |  |  | 0.026 |
| No | 5549 (34) | 4336 (34) | 1213 (36) |  |
| Yes | 10571 (66) | 8420 (66) | 2151 (64) |  |
| **Scoring system** |  |  |  |  |
| SOFA(score) | 6 (4, 8) | 5 (3, 8) | 6 (4, 9) | < 0.001 |
| SAPSII (score) | 38 (30, 49) | 38 (29, 48) | 41 (33, 51) | < 0.001 |
| GCS (score) | 15 (13, 15) | 15 (14, 15) | 14 (11, 15) | < 0.001 |
| **Vital signs** |  |  |  |  |
| HR(minute) | 85.24 (75.32, 97.45) | 85.04 (75.28, 97.13) | 85.91 (75.54, 98.62) | 0.006 |
| SBP(mmHg) | 111.18 (102.27, 123.36) | 110.88 (102.1, 122.92) | 112.67 (102.82, 125.47) | < 0.001 |
| DBP(mmHg) | 62.67 (56.14, 70) | 62.43 (56, 69.68) | 63.67 (57, 71) | < 0.001 |
| MAP(mmHg) | 75 (68.67, 82.77) | 74.75 (68.41, 82.35) | 76 (69.7, 84.34) | < 0.001 |
| RR(minute) | 19.15 (16.85, 22.2) | 19.04 (16.77, 22.13) | 19.45 (17.07, 22.46) | < 0.001 |
| SPO2(%) | 97.17 (95.68, 98.5) | 97.17 (95.68, 98.5) | 97.14 (95.68, 98.52) | 0.645 |
| Temperature(°C ) | 36.87 (36.66, 37.17) | 36.86 (36.65, 37.15) | 36.93 (36.68, 37.28) | < 0.001 |
| ICU_Day(days) | 3.44 (1.93, 7.06) | 3.14 (1.82, 6.39) | 4.9 (2.59, 9.71) | < 0.001 |
| **Laboratory Results** |  |  |  |  |
| RBC(10<sup>12</sup>/L) | 3.43 (2.97, 3.95) | 3.43 (2.98, 3.94) | 3.41 (2.92, 3.97) | 0.089 |
| WBC(10<sup>9</sup>/L) | 12 (8.6, 16.2) | 12.05 (8.68, 16.2) | 11.8 (8.35, 16.3) | 0.18 |
| Platelet(10<sup>9</sup>/L) | 171 (121.24, 237.37) | 169 (121, 235.08) | 178 (122.19, 246) | < 0.001 |
| Hemoglobin(g/dL) | 10.23 (8.81, 11.75) | 10.25 (8.87, 11.73) | 10.15 (8.67, 11.8) | 0.093 |
| RDW(%) | 14.66 (13.5, 16.47) | 14.6 (13.45, 16.3) | 14.97 (13.7, 17) | < 0.001 |
| HCT(%) | 31.3 (27.3, 35.78) | 31.3 (27.34, 35.7) | 31.3 (27.1, 36.14) | 0.573 |
| MCH(pg picograms) | 30.15 (28.58, 31.58) | 30.15 (28.6, 31.53) | 30.1 (28.5, 31.77) | 0.537 |
| MCHC(g/dL) | 32.75 (31.67, 33.75) | 32.8 (31.73, 33.8) | 32.5 (31.43, 33.53) | < 0.001 |
| MCV(fL femtoliters) | 91.67 (87.67, 96) | 91.5 (87.5, 95.67) | 92.33 (88, 97) | < 0.001 |
| INR(ratio) | 1.3 (1.2, 1.58) | 1.3 (1.2, 1.57) | 1.3 (1.15, 1.6) | 0.842 |
| PT(seconds) | 14.3 (12.7, 17.1) | 14.3 (12.75, 17) | 14.3 (12.5, 17.5) | 0.763 |
| APTT(seconds) | 31.68 (27.9, 39.5) | 31.75 (28, 39.5) | 31.38 (27.62, 39.39) | 0.024 |
| Creatinine(mg/dL) | 1.05 (0.77, 1.7) | 1.05 (0.75, 1.67) | 1.1 (0.77, 1.97) | < 0.001 |
| BUN(mg/dL) | 21 (14, 35.67) | 20.5 (14, 34.5) | 23.5 (14.5, 40) | < 0.001 |
| Anion Gap(mEq/L) | 14 (12, 16.67) | 14 (11.67, 16.5) | 14.25 (12, 17) | < 0.001 |
| PH | 7.37 (7.33, 7.41) | 7.37 (7.33, 7.41) | 7.37 (7.32, 7.41) | 0.066 |
| Bicarbonate(mEq/L) | 22.25 (19.67, 24.5) | 22.33 (19.67, 24.5) | 22 (19.33, 25) | 0.213 |
| Calcium(mg/dL) | 8.23 (7.8, 8.7) | 8.2 (7.8, 8.7) | 8.3 (7.85, 8.8) | < 0.001 |
| Magnesium(mg/dL) | 2.03 (1.85, 2.27) | 2.04 (1.85, 2.28) | 2.02 (1.85, 2.25) | 0.005 |
| Chloride(mEq/L) | 104.4 (100, 108) | 104.5 (100.5, 108) | 104 (99.33, 107.5) | < 0.001 |
| Potassium(mEq/L) | 4.18 (3.85, 4.56) | 4.2 (3.85, 4.55) | 4.13 (3.8, 4.6) | 0.008 |
| Sodium(mEq/L) | 138.33 (135.5, 141) | 138 (135.5, 140.5) | 139 (135.67, 141.87) | < 0.001 |
| Lactate(mmol/L) | 1.83 (1.33, 2.6) | 1.85 (1.35, 2.61) | 1.8 (1.3, 2.6) | < 0.001 |
| Glucose(mg/dL) | 129 (108.33, 162.5) | 128.54 (108.33, | 131 (108, 166.5) | 0.02 |

SOFA, Sequential Organ Failure Assessment; SAPSII, Simplified Acute Physiologic Score II; GCS, Glasgow Coma Scale; SBP, Systolic Blood Pressure; DBP, Diastolic Blood Pressure; MAP, Mean Arterial Pressure; RDW, Red Cell Distribution Width; MCHC, Mean Corpuscular Hemoglobin Concentration; MCV, Mean Corpuscular Volume; BUN, Blood Urea Nitrogen; AKI, Acute Kidney Injury; T2DM, Type 2 Diabetes Mellitus; HF, Heart Failure; MV, Mechanical Ventilation; RBC, Red Blood Cell; WBC, White Blood Cell; HCT, Hematocrit; MCH, Mean Corpuscular Hemoglobin; INR, International Normalized Ratio; PT, Prothrombin Time; APTT, Activated Partial Thromboplastin Time.

### 3.2 Variable Selection

The 16120 patients collected from the MIMIC-IV database were randomly divided into a training set (70%) and an internal validation set (30%). To exclude irrelevant variables, we performed an initial screening of variables using MLR, Lasso regression, and the Boruta algorithm. As shown in Table 2, ULR analysis indicated statistical relationships between AKI, Type 2 Diabetes Mellitus, Heart Failure, MV, CRRT, Midazolam, VP, Age, Weight, ICU Day, SOFA, SAPS II, GCS, SBP, DBP, MAP, Temperature, Platelet, RDW, MCHC, MCV, Creatinine, Blood Urea Nitrogen, Anion Gap, Calcium, Magnesium, Chloride, Sodium, Lactate, and Glucose with SAD patients (P < 0.05). MLR analysis of these results suggested that these factors have significant statistical associations with SAD patients, indicating they may be independent risk factors for patient mortality. The best Lasso regression lambda.1se value was confirmed to be 0.0035 through 20-fold cross-validation, resulting in the identification of 28 variables with significant predictive power (Fig 3A, B). Additionally, we utilized the Boruta algorithm for a more in-depth analysis to clarify key variables. This algorithm effectively distinguishes between strongly correlated and weakly correlated variables, significantly improving predictive accuracy. In the analysis results presentation, the green boxes indicate shadow features automatically generated by the algorithm, which were excluded from the final analysis to focus on the most influential variables. Ultimately, we identified 40 impactful variables (Fig 3C, D). Finally, we generated a Venn diagram using the “ ggvenn ” package in R, revealing 17 features shared among the three algorithms (Fig 3E). Based on these features, we established eight machine learning predictive models.

**Fig 3.**
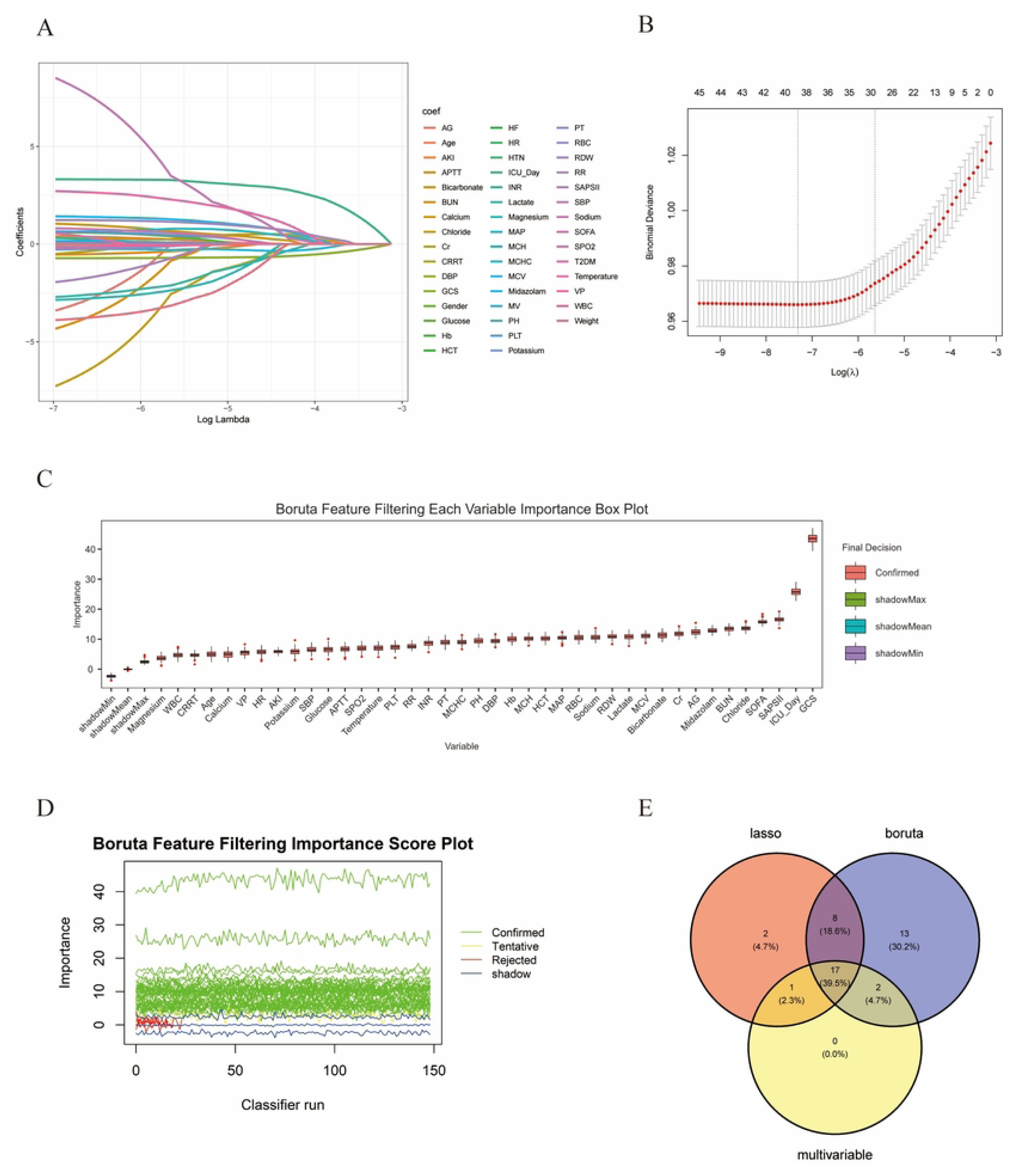
Features identified through Lasso analyses and Boruta. (A) Variable Trajectory Screening via LASSO Regression. (B) The LASSO model was subjected to 20-fold cross-validation to determine the optimal penalization parameter (lambda.1se). (C, D) Variables selected by the boruta algorithm. In terms of feature importance scores, the 40 red variables are considered to be important variables. (E) The Venn diagram illustrates the features selected by Boruta, Lasso, and MLR. The intersection of the features identified by these three methods reveals 17 clinical characteristics.

**Table 2.**
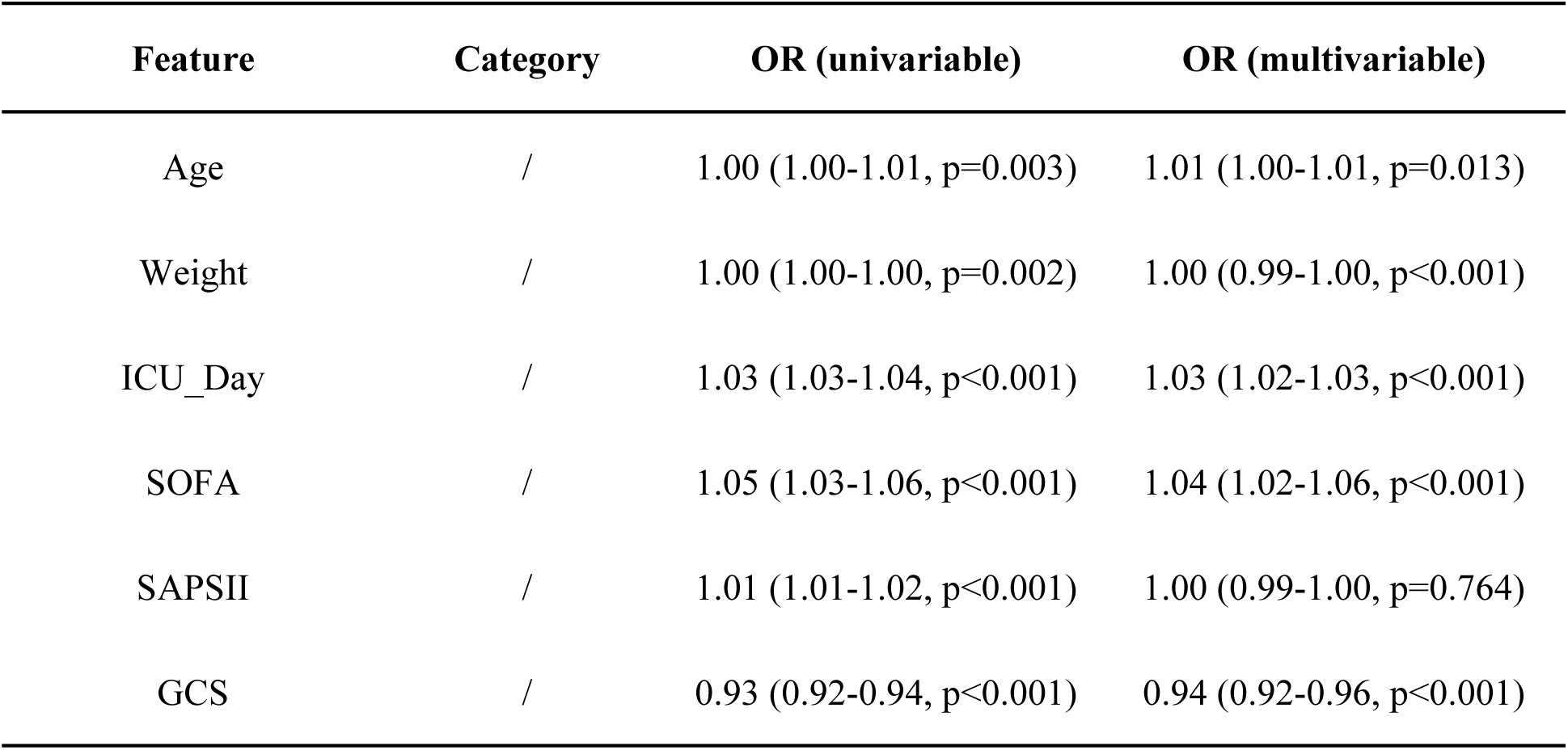
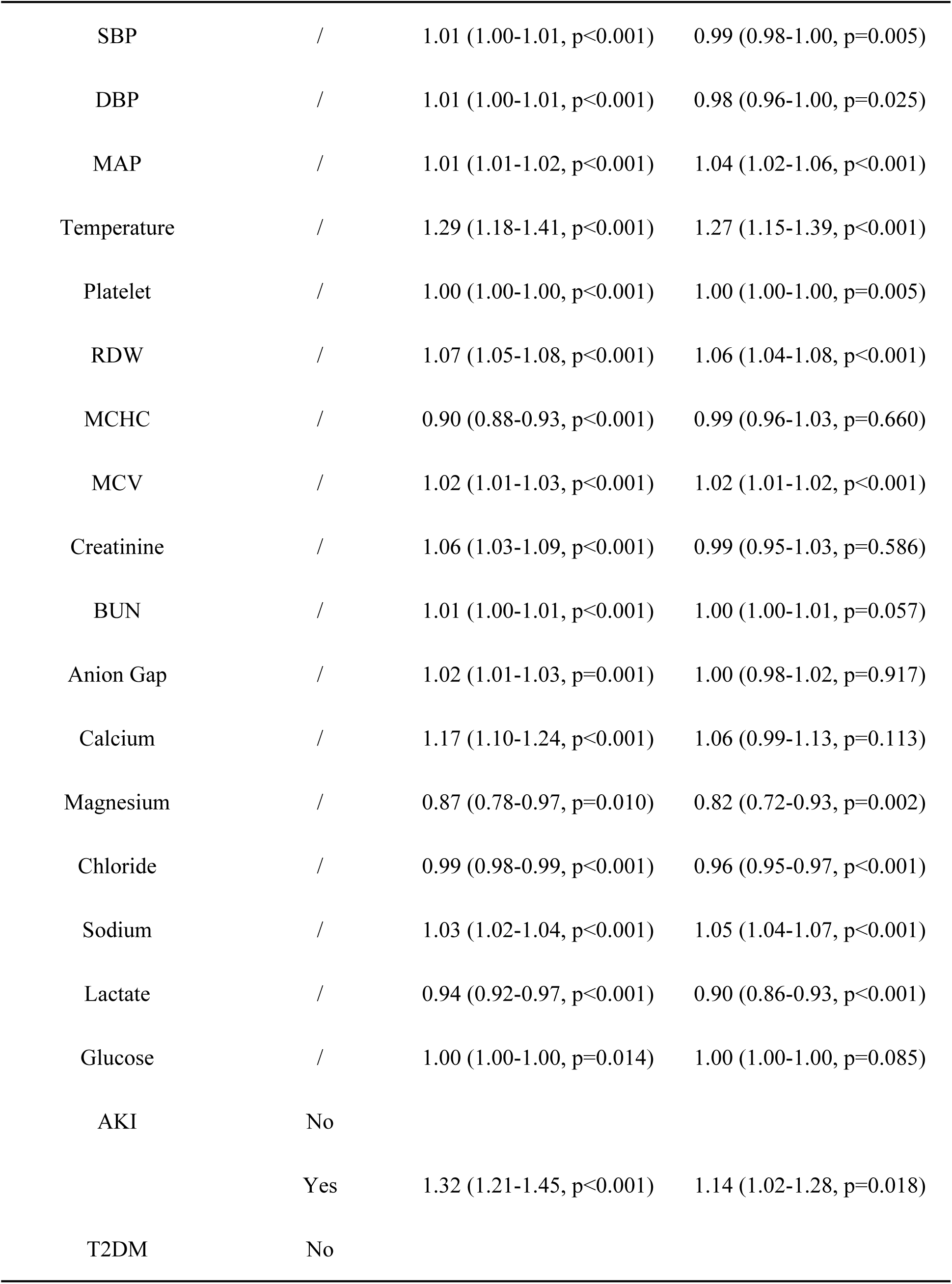
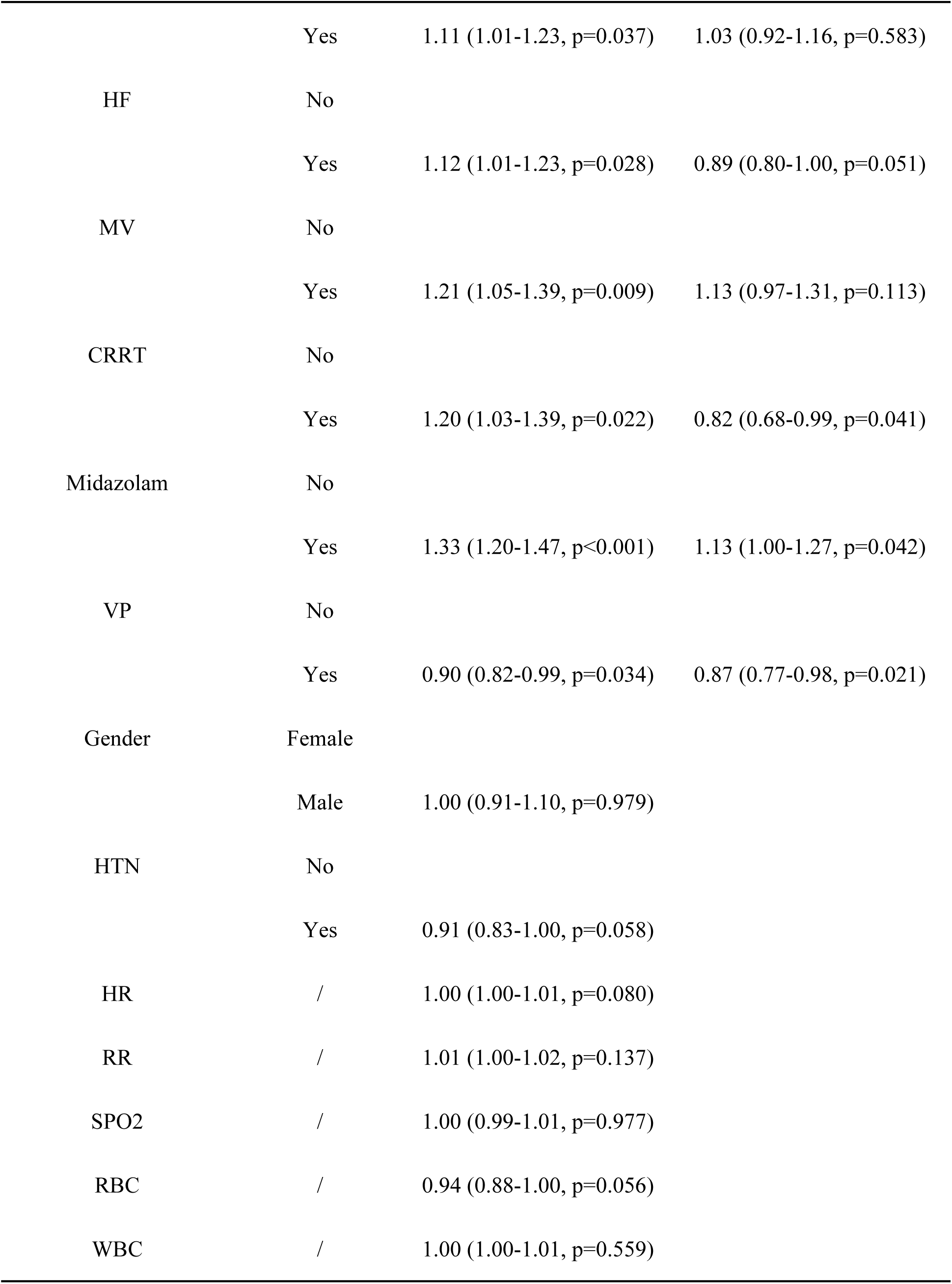
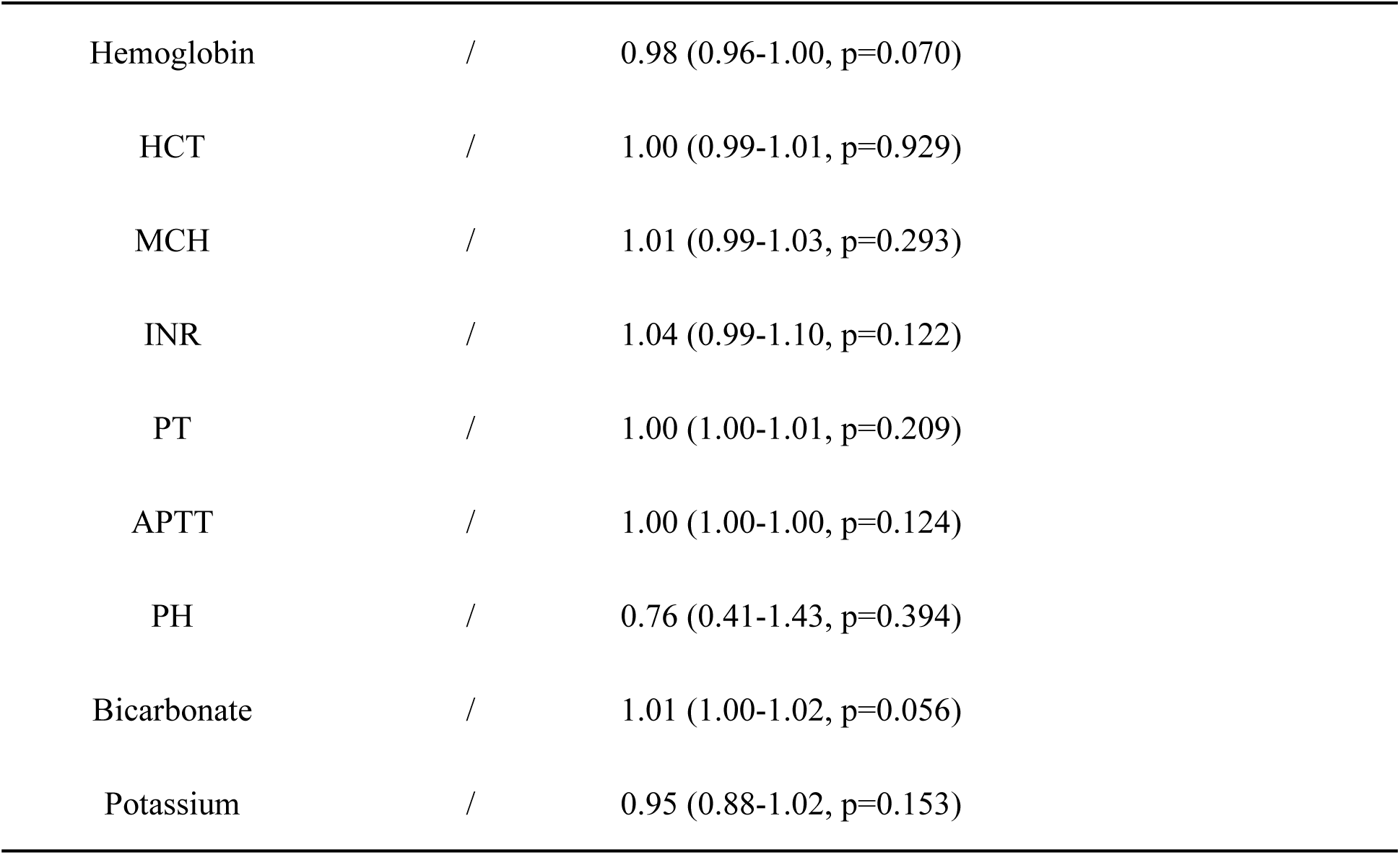
Results from univariate and multivariate logistic regression analyses conducted on the training set.

| Feature | Category | OR (univariable) | OR (multivariable) |
| --- | --- | --- | --- |
| Age | / | 1.00 (1.00-1.01, p=0.003) | 1.01 (1.00-1.01, p=0.013) |
| Weight | / | 1.00 (1.00-1.00, p=0.002) | 1.00 (0.99-1.00, p<0.001) |
| ICU_Day | / | 1.03 (1.03-1.04, p<0.001) | 1.03 (1.02-1.03, p<0.001) |
| SOFA | / | 1.05 (1.03-1.06, p<0.001) | 1.04 (1.02-1.06, p<0.001) |
| SAPSII | / | 1.01 (1.01-1.02, p<0.001) | 1.00 (0.99-1.00, p=0.764) |
| GCS | / | 0.93 (0.92-0.94, p<0.001) | 0.94 (0.92-0.96, p<0.001) |
| SBP | / | 1.01 (1.00-1.01, p<0.001) | 0.99 (0.98-1.00, p=0.005) |
| DBP | / | 1.01 (1.00-1.01, p<0.001) | 0.98 (0.96-1.00, p=0.025) |
| MAP | / | 1.01 (1.01-1.02, p<0.001) | 1.04 (1.02-1.06, p<0.001) |
| Temperature | / | 1.29 (1.18-1.41, p<0.001) | 1.27 (1.15-1.39, p<0.001) |
| Platelet | / | 1.00 (1.00-1.00, p<0.001) | 1.00 (1.00-1.00, p=0.005) |
| RDW | / | 1.07 (1.05-1.08, p<0.001) | 1.06 (1.04-1.08, p<0.001) |
| MCHC | / | 0.90 (0.88-0.93, p<0.001) | 0.99 (0.96-1.03, p=0.660) |
| MCV | / | 1.02 (1.01-1.03, p<0.001) | 1.02 (1.01-1.02, p<0.001) |
| Creatinine | / | 1.06 (1.03-1.09, p<0.001) | 0.99 (0.95-1.03, p=0.586) |
| BUN | / | 1.01 (1.00-1.01, p<0.001) | 1.00 (1.00-1.01, p=0.057) |
| Anion Gap | / | 1.02 (1.01-1.03, p=0.001) | 1.00 (0.98-1.02, p=0.917) |
| Calcium | / | 1.17 (1.10-1.24, p<0.001) | 1.06 (0.99-1.13, p=0.113) |
| Magnesium | / | 0.87 (0.78-0.97, p=0.010) | 0.82 (0.72-0.93, p=0.002) |
| Chloride | / | 0.99 (0.98-0.99, p<0.001) | 0.96 (0.95-0.97, p<0.001) |
| Sodium | / | 1.03 (1.02-1.04, p<0.001) | 1.05 (1.04-1.07, p<0.001) |
| Lactate | / | 0.94 (0.92-0.97, p<0.001) | 0.90 (0.86-0.93, p<0.001) |
| Glucose | / | 1.00 (1.00-1.00, p=0.014) | 1.00 (1.00-1.00, p=0.085) |
| AKI | No |  |  |
|  | Yes | 1.32 (1.21-1.45, p<0.001) | 1.14 (1.02-1.28, p=0.018) |
| T2DM | No |  |  |
|  | Yes | 1.11 (1.01-1.23, p=0.037) | 1.03 (0.92-1.16, p=0.583) |
| HF | No |  |  |
|  | Yes | 1.12 (1.01-1.23, p=0.028) | 0.89 (0.80-1.00, p=0.051) |
| MV | No |  |  |
|  | Yes | 1.21 (1.05-1.39, p=0.009) | 1.13 (0.97-1.31, p=0.113) |
| CRRT | No |  |  |
|  | Yes | 1.20 (1.03-1.39, p=0.022) | 0.82 (0.68-0.99, p=0.041) |
| Midazolam | No |  |  |
|  | Yes | 1.33 (1.20-1.47, p<0.001) | 1.13 (1.00-1.27, p=0.042) |
| VP | No |  |  |
|  | Yes | 0.90 (0.82-0.99, p=0.034) | 0.87 (0.77-0.98, p=0.021) |
| Gender | Female |  |  |
|  | Male | 1.00 (0.91-1.10, p=0.979) |  |
| HTN | No |  |  |
|  | Yes | 0.91 (0.83-1.00, p=0.058) |  |
| HR | / | 1.00 (1.00-1.01, p=0.080) |  |
| RR | / | 1.01 (1.00-1.02, p=0.137) |  |
| SPO2 | / | 1.00 (0.99-1.01, p=0.977) |  |
| RBC | / | 0.94 (0.88-1.00, p=0.056) |  |
| WBC | / | 1.00 (1.00-1.01, p=0.559) |  |

|  |  |  |
| --- | --- | --- |
| Hemoglobin | / | 0.98 (0.96-1.00, p=0.070) |
| HCT | / | 1.00 (0.99-1.01, p=0.929) |
| MCH | / | 1.01 (0.99-1.03, p=0.293) |
| INR | / | 1.04 (0.99-1.10, p=0.122) |
| PT | / | 1.00 (1.00-1.01, p=0.209) |
| APTT | / | 1.00 (1.00-1.00, p=0.124) |
| PH | / | 0.76 (0.41-1.43, p=0.394) |
| Bicarbonate | / | 1.01 (1.00-1.02, p=0.056) |
| Potassium | / | 0.95 (0.88-1.02, p=0.153) |

SOFA, Sequential Organ Failure Assessment; SAPSII, Simplified Acute Physiologic Score II; GCS, Glasgow Coma Scale; SBP, Systolic Blood Pressure; DBP, Diastolic Blood Pressure; MAP, Mean Arterial Pressure; RDW, Red Cell Distribution Width; MCHC, Mean Corpuscular Hemoglobin Concentration; MCV, Mean Corpuscular Volume; BUN, Blood Urea Nitrogen; AKI, Acute Kidney Injury; T2DM, Type 2 Diabetes Mellitus; HF, Heart Failure; MV, Mechanical Ventilation; RBC, Red Blood Cell; WBC, White Blood Cell; HCT, Hematocrit; MCH, Mean Corpuscular Hemoglobin; INR, International Normalized Ratio; PT, Prothrombin Time; APTT, Activated Partial Thromboplastin Time.

### 3.3 Model performance on test and external validation datasets

After identifying 17 clinical features, we constructed predictive models for SAD using eight machine learning methods. After applying the SMOTE function to balance the training set, we developed models based on LR, SVM, GBM, Neural Network, RF, XGBoost, AdaBoost, and LightGBM, yielding AUC values of 0.661, 0.657, 0.831, 0.705, 1.00, 0.733, 0.695, and 0.789, respectively (Fig 4A). In the internal validation set, the AUC values were 0.670, 0.677, 0.730, 0.702, 0.732, 0.714, 0.670, and 0.713 (Fig 4B). Table 3 provides the Accuracy, Sensitivity, Specificity, Precision, and F1-Score for both the training and internal validation sets. Due to the substantial discrepancy between the training and validation sets for the RF model, raising concerns about potential overfitting, we ultimately selected GBM as the best model. To further evaluate the performance of the GBM model, we plotted the calibration curves for both the training and internal validation sets, and also generated the Decision Curve Analysis (DCA) curves for both sets. The calibration curves and DCA for the GBM model demonstrate good predictive performance and significant clinical net benefits across most threshold probability ranges (Fig 4C, F). Based on these performance metrics, the GBM model was confirmed as the most suitable predictive tool for this dataset and was selected for subsequent analyses. The confusion matrix presents the classification results of the GBM model for the patients in the internal validation set (Fig 4G). Finally, we also illustrated the feature selection for the GBM model (Fig 4H).

**Fig 4.**
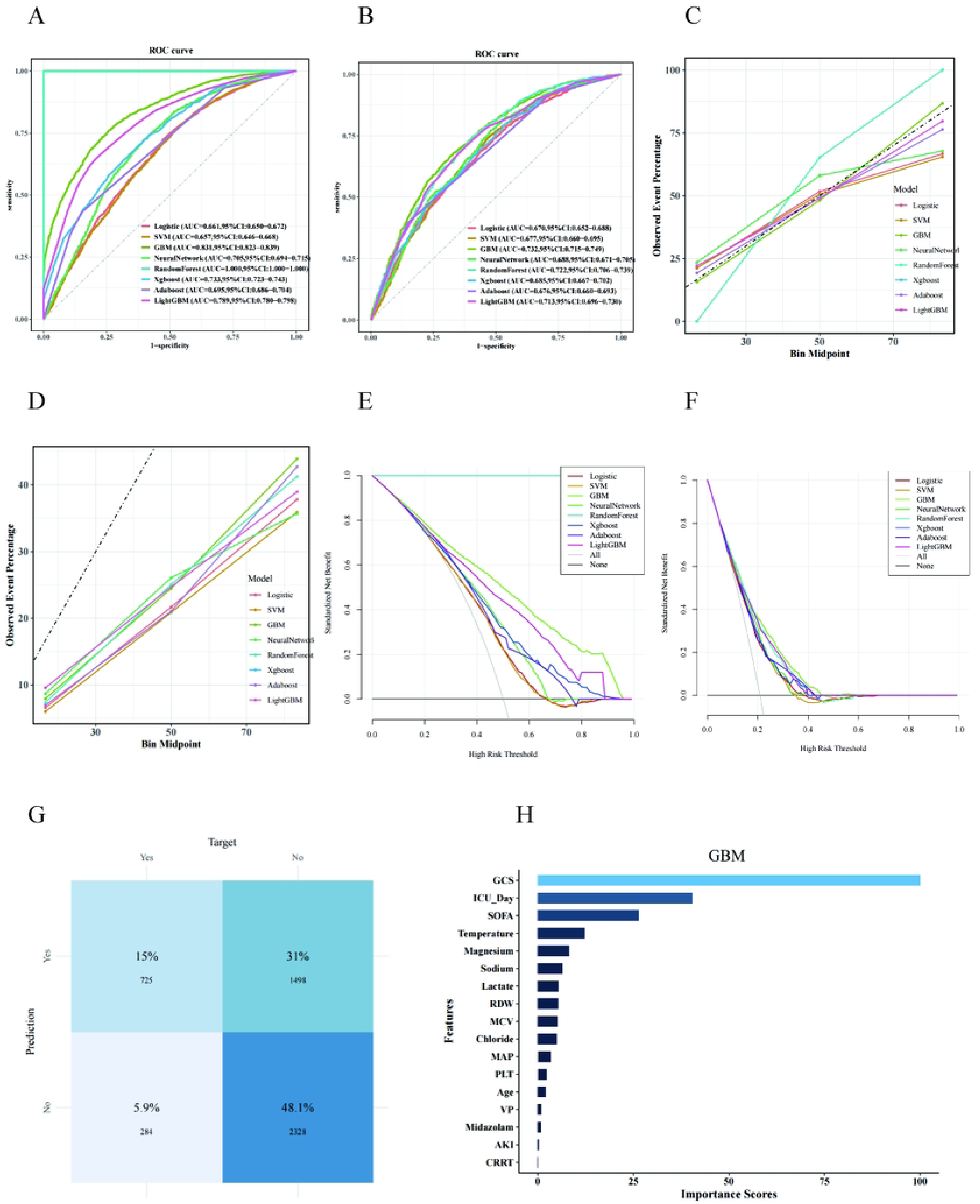
A comparative analysis of the performance of eight distinct predictive models. (A) The ROC curve of the training set. (B) The ROC curve of the internal validation set. (C, D) The calibration curves of the training set and internal validation set. (E, F) The clinical decision curves of the internal validation set and external validation set. (G) The confusion matrix of the internal validation set. (H) Feature Selection for the GBM Model.

**Table 3.** The predictive capabilities of each model.

| Model | Accuracy | Sensitivity | Specificity | Precision | F1 |
| --- | --- | --- | --- | --- | --- |
| Train sets |  |  |  |  |  |
| Logistic | 0.626 | 0.725 | 0.527 | 0.605 | 0.66 |
| SVM | 0.621 | 0.78 | 0.462 | 0.592 | 0.673 |
| GBM | 0.751 | 0.734 | 0.768 | 0.76 | 0.747 |
| NeuralNetwork | 0.663 | 0.72 | 0.607 | 0.647 | 0.681 |
| RandomForest | 1 | 1 | 1 | 1 | 1 |
| Xgboost | 0.663 | 0.742 | 0.585 | 0.641 | 0.688 |
| Adaboost | 0.64 | 0.43 | 0.85 | 0.741 | 0.544 |
| LightGBM | 0.72 | 0.655 | 0.784 | 0.752 | 0.7 |
| Internal validation sets |  |  |  |  |  |
| Logistic | 0.589 | 0.687 | 0.563 | 0.293 | 0.411 |
| SVM | 0.576 | 0.742 | 0.532 | 0.295 | 0.422 |
| GBM | 0.681 | 0.665 | 0.685 | 0.358 | 0.465 |
| NeuralNetwork | 0.574 | 0.77 | 0.522 | 0.298 | 0.43 |
| RandomForest | 0.625 | 0.747 | 0.593 | 0.326 | 0.454 |
| Xgboost | 0.535 | 0.796 | 0.466 | 0.282 | 0.417 |
| Adaboost | 0.756 | 0.395 | 0.852 | 0.413 | 0.404 |
| LightGBM | 0.631 | 0.719 | 0.608 | 0.326 | 0.449 |
| Train sets |  |  |  |  |  |
| Logistic | 0.626 | 0.725 | 0.527 | 0.605 | 0.66 |
| SVM | 0.621 | 0.78 | 0.462 | 0.592 | 0.673 |

### 3.4 Interpretability Analysis

To further explore the clinical application of the GBM model, we utilized the SHAP algorithm to quantify the contribution of each feature within the model. The feature importance bee swarm plot we created illustrates the mechanisms by which each feature affects the model’s predictions, with SHAP values plotted on the x-axis; higher SHAP values indicate an increased likelihood of the outcome occurring, and vice versa. The y-axis represents the magnitude of the feature values, visually depicted through a gradient from yellow to purple, where yellow indicates high feature values and purple signifies low feature values. According to the results, lower GCS scores, lower Chloride levels, and higher ICU Day, SOFA scores, Sodium levels, RDW, MCV, Age, and the administration of Midazolam are associated with higher SHAP values, indicating a greater likelihood of delirium in sepsis patients (Fig 5A). Fig 5B displays the GBM model’s SHAP significance analysis, visualizing the ranking of feature importance. To enhance the interpretability of the model’s decision-making process at the individual level, we conducted a systematic interpretability study on two representative cases. We plotted bar charts for one SAD patient and one Non-SAD patient, where yellow represents increased risk and purple indicates reduced risk (Fig 5C, D). By visualizing the SHAP values for these samples, we can intuitively identify the influence of each feature on the model’s predictions in specific instances. Furthermore, to explore the interactions between different variables, we used SOFA scores as an example; the SHAP values corresponding to GCS, Sodium, and Lactate levels may vary with different SOFA scores (Fig 6A, C).

**Fig 5.**
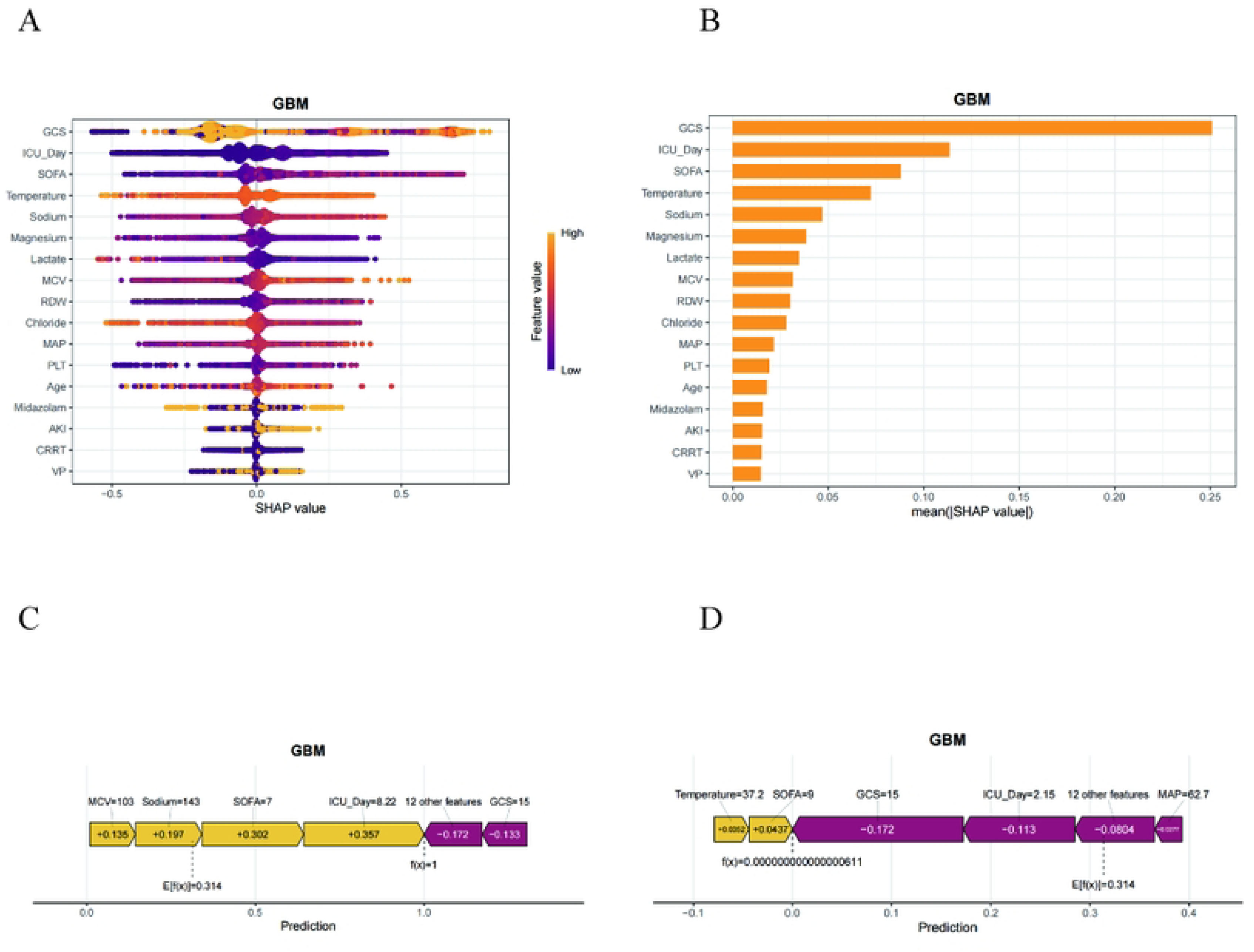
SHAP analysis of the GBM model. (A) The SHAP summary plot of the GBM model. (B) Significance analysis of feature importance ranking via SHAP, based on the mean value. (C, D) The force plots offer individualized feature attributions for two representative examples.C: Patients with SAD; D: Patients with Non-SAD.

**Fig 6.**
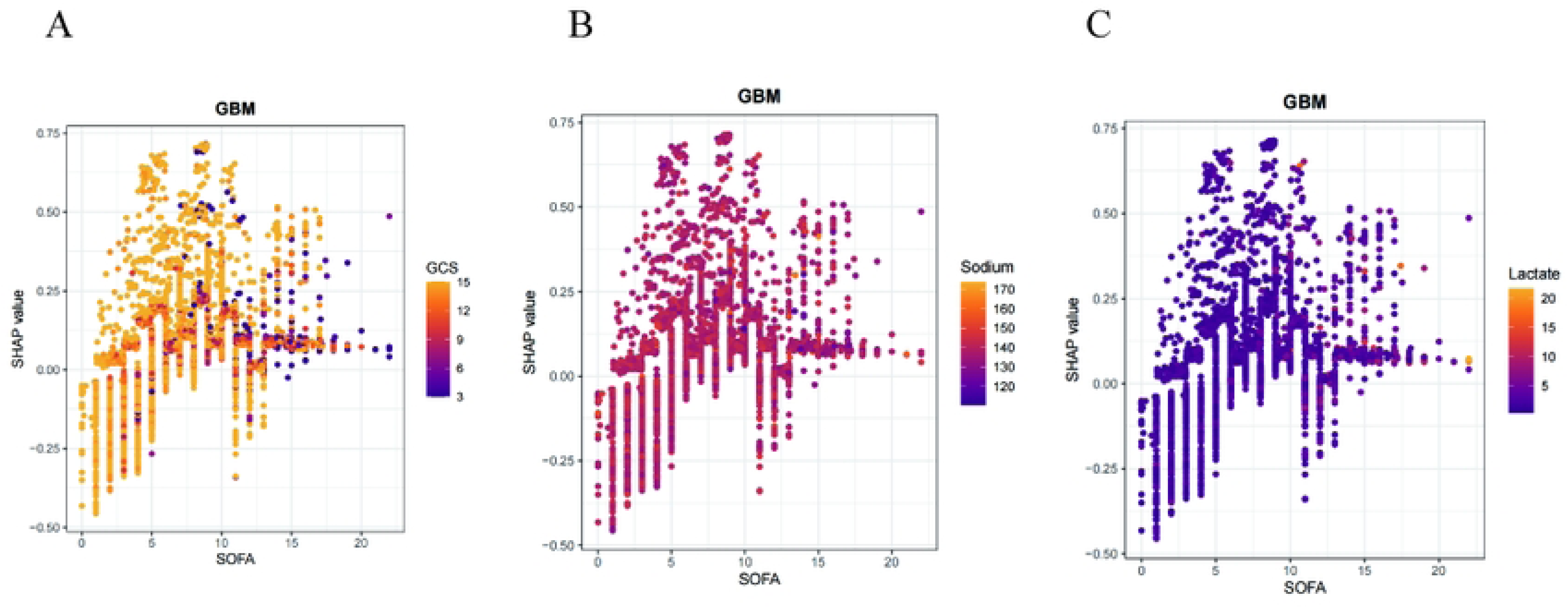
The SHAP dependence plot for features in the GBM model. (A-C) The Y-axis represents SHAP values, while the X-axis depicts actual clinical parameters.It is noteworthy that when the SHAP value of a feature is greater than 0, it indicates an increased risk of SAD, while a negative SHAP value suggests a reduced risk.

### 3.5 Construction of the Online Calculator

In this study, we developed an online web calculator based on the GBM model (Fig 7A) (https://risk-model.shinyapps.io/make_web/). This calculator can predict the likelihood of delirium occurring in sepsis patients within the ICU based on various clinical feature variables.

**Fig 7.**
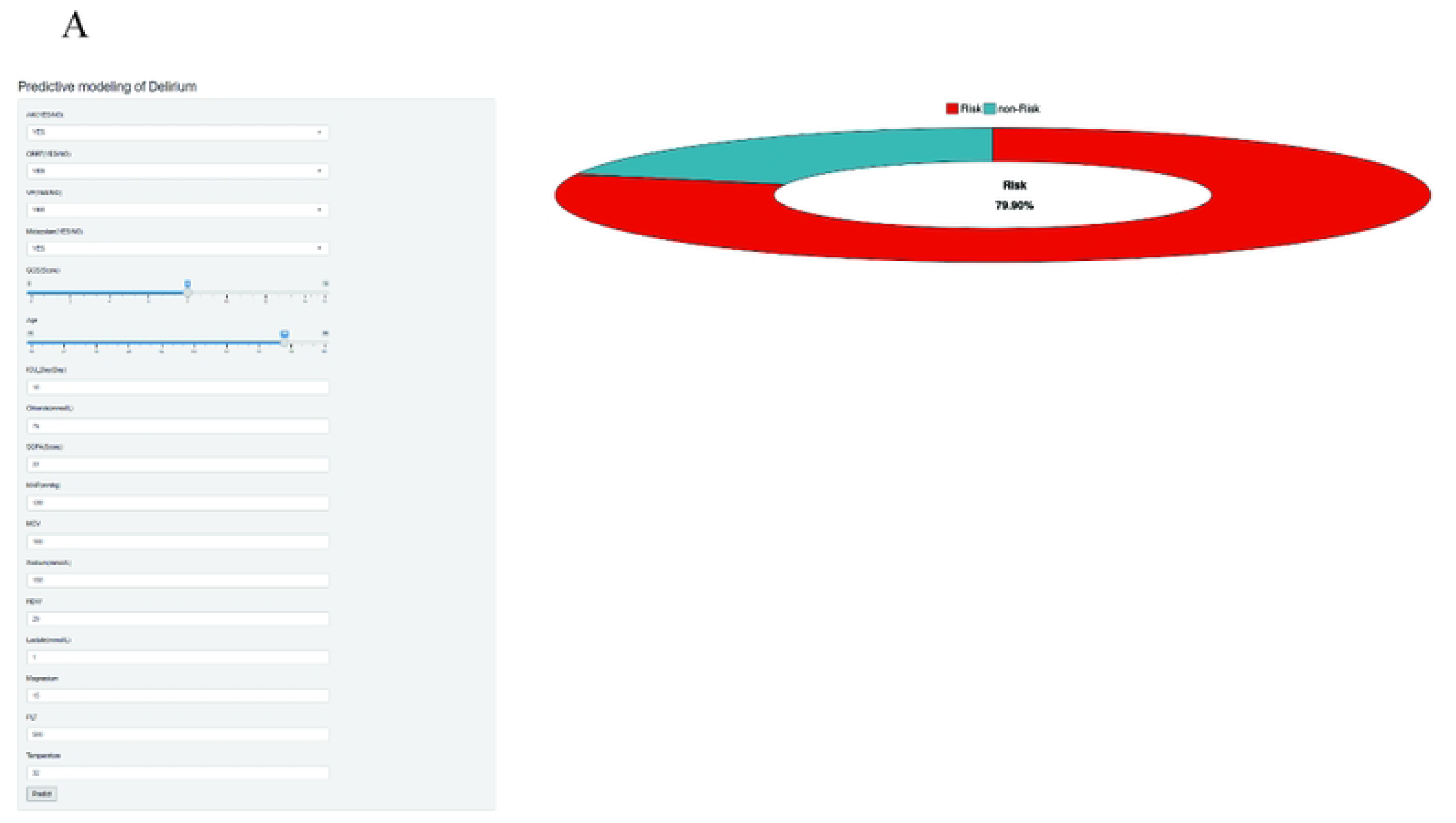
A Online **web calculator for predicting the probability of SAD occurrence.**

Clinicians can conveniently input relevant data into this tool, facilitating the use of the model to predict the incidence of SAD and allowing for timely adjustments to treatment plans to improve patient outcomes.

## 4. Discussion

SAD is a severe neurological syndrome that significantly increases mortality among affected patients, leading to long-term mental and cognitive impairments, and even dementia, placing a substantial burden on patients and their families (21, 22). We developed a machine learning prediction model based on GBM, which showed promising results in internal validation (AUC: 0.732). Utilizing the SHAP method, we enhanced the clinical interpretability of the model. Although several predictive models have been developed to assess the risk of SAD in the ICU, their practicality in clinical practice remains insufficient, failing to effectively translate into clinical tools (6, 7). Therefore, we created a simple web-based calculator to assist healthcare professionals in quickly identifying SAD and timely adjusting medical strategies to improve patient outcomes.

Currently, clinical recognition of SAD in the ICU is not optimistic, mainly due to physicians’ insufficient understanding of the complication. This underscores the importance of enhancing clinical awareness. Through systematic training, we can improve healthcare teams’ ability to recognize sepsis-associated delirium, thereby allowing for more accurate identification of early intervention opportunities and a scientific assessment of the risk-benefit ratio of treatment strategies. Commonly used delirium assessment methods in the ICU include CAM-ICU and the Intensive Care Delirium Screening Checklist (ICDSC). CAM-ICU is applicable for patients requiring mechanical ventilation, offering rapid and effective assessment; however, it is not suitable for deeply sedated or comatose patients and requires multiple evaluations for diagnosis (23–25). In contrast, the ICDSC has a broader applicability, as it does not require patient cooperation and is based on nurses observing patients’ behavior over a 24-hour period. Nonetheless, it relies on nurses’ subjective judgment, which may lead to lower consistency among different evaluators and increase the nursing workload (26–28). The time-consuming nature of both methods may delay treatment decisions and compromise patient safety. To address this issue, we constructed a web-based calculator using the GBM model to enable clinicians to quickly detect SAD early. It is important to emphasize that, although preliminary validation shows promising performance, multi-center, prospective cohort studies are still required to ensure the model’s applicability in various clinical scenarios, providing external validation with independent datasets. This will help objectively assess the model’s generalizability and diagnostic accuracy, offering evidence-based medicine to support its clinical translation.

Feature selection is a crucial step in building predictive models, as its validity directly affects the model’s predictive performance (29). We obtained a substantial sample from the MIMIC-IV database and utilized MLR to identify independent risk factors for SAD patients. Lasso regression was applied to process features, avoiding multicollinearity and reducing the risk of overfitting (30). Additionally, we utilized the Boruta algorithm for variable selection, employing a rigorous statistical significance filtering mechanism to retain key features while suppressing model complexity and enhancing the model’s generalizability on independent datasets. Ultimately, we included common variables to achieve accurate feature selection and stability. In the realm of healthcare big data, machine learning algorithms demonstrate significant advantages in handling complex medical data (31). Compared to traditional statistical models, the nonlinear modeling capabilities of machine learning can capture higher-order interactions between variables and enable multivariable synchronous analysis of high-dimensional data through parallel computing frameworks, thereby increasing the ability to predict diseases (32). In this study, we applied eight machine learning models for performance comparison and ultimately chose GBM as the best model.

GBM is one of the most representative methods in ensemble learning, which iteratively combines weak learners and utilizes gradient descent to optimize prediction errors, simulating the progressive learning and collaborative optimization characteristics found in biological systems (33–35). As an efficient predictive model, GBM is able to capture nonlinear relationships and complex interaction effects in the data and is widely used in the construction of clinical prediction models. The clinical interpretability of machine learning models is critical for medical practice; however, interpretability has long been one of the core challenges in this field (36). To address this issue, we adopted the SHAP method to analyze features and enhance model interpretability (37). Compared to traditional weight-based explanatory methods, SHAP exhibits superior consistency and performance, while demonstrating greater stability across various models (38). This study utilized SHAP value analysis, which significantly improved the model’s interpretability compared to the coefficient interpretations of traditional generalized linear regression models. SHAP values not only quantify the contributions of each feature to the predictive outcomes but also provide intuitive visualizations through feature importance plots (39). This analytical approach offers a new perspective for understanding the decision-making mechanisms of machine learning models, clearly illustrating the specific impacts of feature variables on model predictions, thereby effectively enhancing the model’s interpretability and transparency.

According to the SHAP feature importance plot, a low GCS score is identified as the most significant risk factor for SAD. Studies have shown that low GCS scores, high SOFA scores, advanced age, prolonged ICU stay, hypernatremia, and the use of midazolam are all risk factors for SAD. Specifically, the GCS score is one of the strong predictive features, originally designed by Graham Teasdale and Bryan Jennett at the University of Glasgow for assessing traumatic brain injury. It quantifies eye-opening, verbal, and motor responses to objectively evaluate the degree of consciousness impairment (40). The lower the GCS score, the higher the risk of delirium; research indicates that for each point decrease in GCS, the risk of delirium increases by approximately 34% (41, 42). Notably, SHAP analysis also revealed a complex nonlinear relationship between GCS scores and SAD: scores of 1-3 typically indicate severe brain injury, whereas a score of 15 suggests full consciousness; the probability of delirium occurrence is significantly higher in the former group compared to those in the intermediate scoring range, a phenomenon that has also been reported in other delirium prediction models (43). The SOFA score is an effective tool for assessing organ dysfunction in sepsis patients, with the central nervous system score relying on the GCS. By treating GCS as an independent variable, we avoided the limitation of the SOFA score, which might focus solely on single-organ function assessment, thus more accurately reflecting brain function impairment. An elevated SOFA score signifies systemic inflammatory response, tissue hypoxia, and organ dysfunction, with central nervous system dysfunction being the most common. These factors are interrelated via a systemic inflammation-brain injury axis, where many inflammatory factors can disrupt the blood-brain barrier and subsequently induce delirium (44–46). Research has found that patients with a SOFA score exceeding 9 have a probability of SAD occurrence greater than 70%, making the SOFA score a core indicator for predicting SAD; a high SOFA score is an independent risk factor for delirium (47).

Advanced age and prolonged ICU stay are both significant risk factors for delirium. As age increases, older patients often have various chronic diseases, malnutrition, sensory impairments, and cognitive deficits, resulting in decreased brain physiological reserve, thereby heightening the probability of delirium (48). Patients who are in the ICU for extended periods are more likely to be exposed to mechanical ventilation, sedative medications, sleep deprivation, and infections, which can further trigger delirium. Delirious patients commonly exhibit complications such as agitation, respiratory dysfunction, and infections, which may significantly prolong their ICU stays, creating a vicious cycle. Studies have shown that remaining in the ICU for more than 7 days can transition delirium from an “acute” to a “persistent” state, with about 30%-40% of patients experiencing long-term cognitive sequelae (49, 50). Additionally, midazolam, as a benzodiazepine, suppresses the central nervous system, potentially interfering with cholinergic neurotransmission and disrupting sleep architecture, thus increasing the risk of delirium. Our findings indicate that the pre-illness use of midazolam significantly raises the risk of delirium, consistent with recent studies (51, 52). Hypernatremia also contributes to an increased incidence of delirium. Hypernatremia affects sodium-potassium pump function, leading to abnormal neural excitability and interfering with the release of inhibitory neurotransmitters such as gamma-aminobutyric acid. Furthermore, hypernatremia decreases brain energy metabolism, affecting glucose utilization and ATP production, resulting in insufficient neuronal energy, which further contributes to the onset of delirium (53–55).

In summary, the SHAP method provides significant support for personalized diagnosis and treatment of SAD patients. By quantifying the contributions of various feature variables to predictive outcomes, it enhances the interpretability of the model’s predictions and offers intuitive decision-making support for clinicians. The SHAP values clearly illustrate the impact of clinical features on patient prognosis, aiding physicians in identifying key risk factors and formulating personalized intervention strategies. By integrating GBM and SHAP, this model not only improves predictive accuracy but also significantly enhances interpretability. The visualization of SHAP values allows clinicians to gain deeper insights into important features and their interactions, thereby increasing the transparency and credibility of medical decisions and providing a scientific basis for the personalized risk management of SAD. This interpretable machine learning approach holds substantial practical value in clinical settings and advances the development of precision medicine.

This study has some limitations. First, as a retrospective analysis, the nature of the study restricts the establishment of causal relationships between features and outcomes. Additionally, certain features (such as PCT and IL-6) had limited data availability in the database, resulting in the exclusion of some potential risk factors from the predictive model. Therefore, future prospective randomized controlled clinical trials should be conducted to validate the model’s effectiveness. Second, the data originated from a single center, lacking sufficient external validation, which may affect the credibility of the GBM model. Lastly, the limitations of sample size reduce the performance of the machine learning algorithms. Future research should establish a systematic data collection mechanism to expand the sample size and diversity, thereby improving the predictive accuracy and reliability of the model and ensuring that the research findings have clinical application value.

## 5. conclusion

In this study, we utilized the MIMIC-IV 3.1 database to develop a GBM model for predicting the risk of SAD. By employing the SHAP method to interpret the intrinsic information of the GBM model, we were able to more intuitively reveal the impact of key features. Our model results indicate that features such as GCS, ICU Day, Chloride, Sodium, SOFA, and Temperature are critical factors in identifying patients at higher risk for delirium. The model demonstrates high predictive accuracy, which is significant for clinical practice and provides important scientific evidence for disease prevention and the formulation of clinical treatment strategies. Furthermore, based on the GBM model, we constructed a web calculator, facilitating the translation of the model into clinical application. This tool can assist healthcare professionals in early and effective prediction of SAD occurrence, helping to optimize management strategies, improve patient outcomes, and enhance the quality of life.

## Data availability statement

This study conducted an in-depth analysis of publicly available datasets, which can be accessed through the following URL: https://mimic.mit.edu.

## Ethics statement

The procedures involving human subjects in this study were approved by the Institutional Review Boards of MIT and Beth Israel Deaconess Medical Center, and the study was conducted in strict accordance with the legal regulations and ethical requirements of the institutions where the research was conducted. During the study, all participants signed written informed consent forms to clearly confirm their agreement to participate. Furthermore, any images or data in the manuscript that may contain personally identifiable information have been granted written permission from the individuals involved, ensuring that the publication of the research findings complies with ethical and legal standards.

## Author contributions

LG conducted this research and drafted the initial manuscript. XYY, SJT, and XJW were responsible for designing the research plan and performing preliminary data analysis. GDW and YXZ revised the manuscript. YRC and JYB participated in the data analysis work. All authors contributed to the completion of the paper and agreed to the submitted version.

## Acknowledgments

The authors sincerely thank the MIMIC official team for their outstanding contributions, which are of significant importance.

## The supplementary table

**S1 Table Hyperparameter settings for eight models.** Gradient Boosting Machine: GBM; Support Vector Machine: SVM; Random Forest: RF; Extreme Gradient Boosting: XGBoost; Adaptive Boosting: AdaBoost; Light Gradient Boosting Machine: LightGBM.

## Abbreviations

Medical Information Mart for Intensive Care IV, MIMIC-IV, sepsis-associated delirium, SAD, Multivariate Logistic Regression, MLR, Least Absolute Shrinkage and Selection Operator Regression, Lasso, The SHapley Additive exPlanations, SHAP, Evaluate the Sequential Organ Failure Assessment, SOFA, Glasgow Coma Scale, GCS, Simplified Acute Physiology Score II, SAPSII, Red Cell Distribution Width, RDW, Mean Corpuscular Hemoglobin, MCH, Mean Corpuscular Hemoglobin Concentration, MCHC, Mean Corpuscular Volume, MCV, Potential of Hydrogen, PH, Acute Kidney Injury, AKI, Mechanical Ventilation, MV, Continuous Renal Replacement Therapy, CRRT, Vasopressin, VP, Confusion Assessment Method for the Intensive Care Unit, CAM-ICU, Intensive Care Delirium Screening Checklist, ICDSC, Logistic Regression, LR, Support Vector Machine, SVM, Gradient Boosting Machine, GBM, Random Forest, RF, Extreme Gradient Boosting, XGBoost, Adaptive Boosting, AdaBoost, Light Gradient Boosting Machine, LightGBM, Structured Query Language, SQL, the Area Under the Receiver Operating Characteristic Curve, AUC, Decision Curve Analysis, DCA

## Reference

1. Tang J, Huang J, He X, Zou S, Gong L, Yuan Q, et al. The prediction of in-hospital mortality in elderly patients with sepsis-associated acute kidney injury utilizing machine learning models. Heliyon. 2024;10(4):e26570.

2. Chung HY, Wickel J, Brunkhorst FM, Geis C. Sepsis-Associated Encephalopathy: From Delirium to Dementia? J Clin Med. 2020;9(3).

3. Fleischmann C, Scherag A, Adhikari NK, Hartog CS, Tsaganos T, Schlattmann P, et al. Assessment of Global Incidence and Mortality of Hospital-treated Sepsis. Current Estimates and Limitations. Am J Respir Crit Care Med. 2016;193(3):259–72.

4. Kempker JA, Martin GS. The Changing Epidemiology and Definitions of Sepsis. Clin Chest Med. 2016;37(2):165–79.

5. Ebersoldt M, Sharshar T, Annane D. Sepsis-associated delirium. Intensive Care Med. 2007;33(6):941–50.

6. Atterton B, Paulino MC, Povoa P, Martin-Loeches I. Sepsis Associated Delirium. Medicina (Kaunas). 2020;56(5).

7. Tokuda R, Nakamura K, Takatani Y, Tanaka C, Kondo Y, Ohbe H, et al. Sepsis-Associated Delirium: A Narrative Review. J Clin Med. 2023;12(4).

8. Greener JG, Kandathil SM, Moffat L, Jones DT. A guide to machine learning for biologists. Nat Rev Mol Cell Biol. 2022;23(1):40–55.

9. Shamout F, Zhu T, Clifton DA. Machine Learning for Clinical Outcome Prediction. IEEE Rev Biomed Eng. 2021;14:116–26.

10. Deo RC. Machine Learning in Medicine. Circulation. 2015;132(20):1920–30.

11. Hunter DJ, Holmes C. Where Medical Statistics Meets Artificial Intelligence. N Engl J Med. 2023;389(13):1211–9.

12. Preliminary criteria for the classification of systemic sclerosis (scleroderma). Subcommittee for scleroderma criteria of the American Rheumatism Association Diagnostic and Therapeutic Criteria Committee. Arthritis Rheum. 1980;23(5):581-90.

13. Vitali C, Bombardieri S, Moutsopoulos HM, Coll J, Gerli R, Hatron PY, et al. Assessment of the European classification criteria for Sjogren’s syndrome in a series of clinically defined cases: results of a prospective multicentre study. The European Study Group on Diagnostic Criteria for Sjogren’s Syndrome. Ann Rheum Dis. 1996;55(2):116–21.

14. Sanchez-Pinto LN, Venable LR, Fahrenbach J, Churpek MM. Comparison of variable selection methods for clinical predictive modeling. Int J Med Inform. 2018;116:10–7.

15. Pate A, Riley RD, Collins GS, van Smeden M, Van Calster B, Ensor J, et al. Minimum sample size for developing a multivariable prediction model using multinomial logistic regression. Stat Methods Med Res. 2023;32(3):555–71.

16. Frost HR, Amos CI. Gene set selection via LASSO penalized regression (SLPR). Nucleic Acids Res. 2017;45(12):e114.

17. Lee S, Gornitz N, Xing EP, Heckerman D, Lippert C. Ensembles of Lasso Screening Rules. IEEE Trans Pattern Anal Mach Intell. 2018;40(12):2841–52.

18. Wang X, Ren J, Ren H, Song W, Qiao Y, Zhao Y, et al. Diabetes mellitus early warning and factor analysis using ensemble Bayesian networks with SMOTE-ENN and Boruta. Sci Rep. 2023;13(1):12718.

19. Dang T, Fermin ASR, Machizawa MG. oFVSD: a Python package of optimized forward variable selection decoder for high-dimensional neuroimaging data. Front Neuroinform. 2023;17:1266713.

20. Poldrack RA, Huckins G, Varoquaux G. Establishment of Best Practices for Evidence for Prediction: A Review. JAMA Psychiatry. 2020;77(5):534–40.

21. Gao Q, Hernandes MS. Sepsis-Associated Encephalopathy and Blood-Brain Barrier Dysfunction. Inflammation. 2021;44(6):2143–50.

22. Sonneville R, Benghanem S, Jeantin L, de Montmollin E, Doman M, Gaudemer A, et al. The spectrum of sepsis-associated encephalopathy: a clinical perspective. Crit Care. 2023;27(1):386.

23. Kotfis K, Marra A, Ely EW. ICU delirium - a diagnostic and therapeutic challenge in the intensive care unit. Anaesthesiol Intensive Ther. 2018;50(2):160–7.

24. Miranda F, Gonzalez F, Plana MN, Zamora J, Quinn TJ, Seron P. Confusion Assessment Method for the Intensive Care Unit (CAM-ICU) for the diagnosis of delirium in adults in critical care settings. Cochrane Database Syst Rev. 2023;11(11):CD013126.

25. Tomasi CD, Grandi C, Salluh J, Soares M, Giombelli VR, Cascaes S, et al. Comparison of CAM-ICU and ICDSC for the detection of delirium in critically ill patients focusing on relevant clinical outcomes. J Crit Care. 2012;27(2):212–7.

26. Fagundes JA, Tomasi CD, Giombelli VR, Alves SC, de Macedo RC, Topanotti MF, et al. CAM-ICU and ICDSC agreement in medical and surgical ICU patients is influenced by disease severity. PLoS One. 2012;7(11):e51010.

27. Krewulak KD, Rosgen BK, Ely EW, Stelfox HT, Fiest KM. The CAM-ICU-7 and ICDSC as measures of delirium severity in critically ill adult patients. PLoS One. 2020;15(11):e0242378.

28. von Hofen-Hohloch J, Awissus C, Fischer MM, Michalski D, Rumpf JJ, Classen J. Delirium Screening in Neurocritical Care and Stroke Unit Patients: A Pilot Study on the Influence of Neurological Deficits on CAM-ICU and ICDSC Outcome. Neurocrit Care. 2020;33(3):708–17.

29. Chowdhury MZI, Turin TC. Variable selection strategies and its importance in clinical prediction modelling. Fam Med Community Health. 2020;8(1):e000262.

30. Tibshirani R. The lasso method for variable selection in the Cox model. Stat Med. 1997;16(4):385–95.

31. Hale AT, Stonko DP, Brown A, Lim J, Voce DJ, Gannon SR, et al. Machine-learning analysis outperforms conventional statistical models and CT classification systems in predicting 6-month outcomes in pediatric patients sustaining traumatic brain injury. Neurosurg Focus. 2018;45(5):E2.

32. Nishi H, Oishi N, Ishii A, Ono I, Ogura T, Sunohara T, et al. Predicting Clinical Outcomes of Large Vessel Occlusion Before Mechanical Thrombectomy Using Machine Learning. Stroke. 2019;50(9):2379–88.

33. Teramoto R. Balanced gradient boosting from imbalanced data for clinical outcome prediction. Stat Appl Genet Mol Biol. 2009;8:Article20.

34. Zeng X. Length of Stay Prediction Model of Indoor Patients Based on Light Gradient Boosting Machine. Comput Intell Neurosci. 2022;2022:9517029.

35. Zhou S, Wang S, Wu Q, Azim R, Li W. Predicting potential miRNA-disease associations by combining gradient boosting decision tree with logistic regression. Comput Biol Chem. 2020;85:107200.

36. Racine AM, Tommet D, D’Aquila ML, Fong TG, Gou Y, Tabloski PA, et al. Machine Learning to Develop and Internally Validate a Predictive Model for Post-operative Delirium in a Prospective, Observational Clinical Cohort Study of Older Surgical Patients. J Gen Intern Med. 2021;36(2):265–73.

37. Song Y, Zhang D, Wang Q, Liu Y, Chen K, Sun J, et al. Prediction models for postoperative delirium in elderly patients with machine-learning algorithms and SHapley Additive exPlanations. Transl Psychiatry. 2024;14(1):57.

38. Gong K, Lee HK, Yu K, Xie X, Li J. A prediction and interpretation framework of acute kidney injury in critical care. J Biomed Inform. 2021;113:103653.

39. Nohara Y, Matsumoto K, Soejima H, Nakashima N. Explanation of machine learning models using shapley additive explanation and application for real data in hospital. Comput Methods Programs Biomed. 2022;214:106584.

40. Rabiu TB. Revisiting the eye opening response of the Glasgow Coma Scale. Indian J Crit Care Med. 2011;15(1):58–9.

41. Bodien YG, Barra A, Temkin NR, Barber J, Foreman B, Vassar M, et al. Diagnosing Level of Consciousness: The Limits of the Glasgow Coma Scale Total Score. J Neurotrauma. 2021;38(23):3295–305.

42. Wang L, Ma X, Zhou G, Gao S, Pan W, Chen J, et al. SOFA in sepsis: with or without GCS. Eur J Med Res. 2024;29(1):296.

43. Zhang Y, Hu J, Hua T, Zhang J, Zhang Z, Yang M. Development of a machine learning-based prediction model for sepsis-associated delirium in the intensive care unit. Sci Rep. 2023;13(1):12697.

44. Myrstad M, Kuwelker K, Haakonsen S, Valebjorg T, Langeland N, Kittang BR, et al. Delirium screening with 4AT in patients aged 65 years and older admitted to the Emergency Department with suspected sepsis: a prospective cohort study. Eur Geriatr Med. 2022;13(1):155–62.

45. Qian X, Sheng Y, Jiang Y, Xu Y. Associations of serum lactate and lactate clearance with delirium in the early stage of ICU: a retrospective cohort study of the MIMIC-IV database. Front Neurol. 2024;15:1371827.

46. Zhang Z, Guo L, Jia L, Duo H, Shen L, Zhao H. Factors contributing to sepsis-associated encephalopathy: a comprehensive systematic review and meta-analysis. Front Med (Lausanne). 2024;11:1379019.

47. Zhao Q, Xiao J, Liu X, Liu H. The nomogram to predict the occurrence of sepsis-associated encephalopathy in elderly patients in the intensive care units: A retrospective cohort study. Front Neurol. 2023;14:1084868.

48. Tang D, Ma C, Xu Y. Interpretable machine learning model for early prediction of delirium in elderly patients following intensive care unit admission: a derivation and validation study. Front Med (Lausanne). 2024;11:1399848.

49. Crimi C, Bigatello LM. The clinical significance of delirium in the intensive care unit. Transl Med UniSa. 2012;2:1–9.

50. Kukreja D, Gunther U, Popp J. Delirium in the elderly: Current problems with increasing geriatric age. Indian J Med Res. 2015;142(6):655–62.

51. Peng W, Shimin S, Hongli W, Yanli Z, Ying Z. Delirium Risk of Dexmedetomidine and Midazolam in Patients Treated with Postoperative Mechanical Ventilation: a Meta-analysis. Open Med (Wars). 2017;12:252–6.

52. Shi HJ, Yuan RX, Zhang JZ, Chen JH, Hu AM. Effect of midazolam on delirium in critically ill patients: a propensity score analysis. J Int Med Res. 2022;50(4):3000605221088695.

53. Ali MA, Hashmi M, Ahmed W, Raza SA, Khan MF, Salim B. Incidence and risk factors of delirium in surgical intensive care unit. Trauma Surg Acute Care Open. 2021;6(1):e000564.

54. Hong L, Shen X, Shi Q, Song X, Chen L, Chen W, et al. Association Between Hypernatremia and Delirium After Cardiac Surgery: A Nested Case-Control Study. Front Cardiovasc Med. 2022;9:828015.

55. Zhao L, Wang Y, Ge Z, Zhu H, Li Y. Mechanical Learning for Prediction of Sepsis-Associated Encephalopathy. Front Comput Neurosci. 2021;15:739265.

